# Understanding the impact of disease and vaccine mechanisms on the importance of optimal vaccine allocation

**DOI:** 10.1101/2022.06.30.22277126

**Authors:** Isobel R. Abell, James M. McCaw, Christopher M. Baker

## Abstract

Vaccination is an important epidemic intervention strategy. Resource limitations and an imperative for efficient use of public resources drives a need for optimal allocation of vaccines within a population. For a disease causing severe illness in particular members of a population, an effective strategy to reduce illness might be to vaccinate those vulnerable with a vaccine that reduces the chance of catching a disease. However, it is not clear that this is the best strategy, and it is generally unclear how the difference between various vaccine strategies changes depending on population characteristics, vaccine mechanisms and allocation objective. In this paper we develop a conceptual mathematical model to consider strategies for vaccine allocation, prior to the establishment of community transmission. By extending the SEIR model to incorporate a range of vaccine mechanisms and disease characteristics, we simulate the impact of vaccination on a population with two sub-groups of differing characteristics. We then compare the outcomes of optimal and suboptimal vaccination strategies for a range of public health objectives using numerical optimisation. Our comparison serves to demonstrate that the difference between vaccinating optimally and suboptimally may be dependent on vaccine mechanism, diseases characteristics, and objective considered. We find that better resources do not guarantee better outcomes. Allocating optimally with lesser vaccine resources can produce a better outcome than allocating good vaccine resources suboptimally, dependent on vaccine mechanisms, disease characteristics and objective considered. Through a principled model-based process, this work highlights the importance of designing effective vaccine allocation strategies. This design process requires models that incorporate known biological characteristics, realistic parameters based on data analysis, etc. Overall, we see that allocation of resources can be just as crucial to the success of a vaccination strategy as the strength of resources available.

## 1. Introduction

Pre-epidemic vaccination is an important intervention for managing disease, and when supply is limited vaccines should be used strategically as part of pandemic preparedness. The difference in the outcomes of various allocation strategies will depend on the characteristics of the pathogen and on vaccine mechanisms. Understanding how these characteristics affect both the optimal allocation strategy and the difference in outcomes between strategies is vital when planning vaccine allocation for novel pathogens or pathogen strains.

When new viruses emerge, decision-makers aim to design effective vaccination strategies based on knowledge of previous pandemics and emerging information [1, 2]. For example, when vaccinating against influenza, there are benefits to targeting vaccination towards those most vulnerable to disease, and targeting vaccination towards those most likely to spread disease [3, 4]. However, these benefits are highly dependent on disease parameters as well as relative efficacy of the vaccine between population groups [5, 6]. When vaccines are available or potential vaccines are in development for novel pathogens, modelling has been used to develop optimal vaccination strategies [7, 8]. However, any strategy developed must be regularly reevaluated as knowledge of pathogen and vaccine characteristics improves and/or new virus variants emerge. For example, the emergence of new variants for COVID-19 in 2021, notably the Delta and Omicron variants, have necessitated the reevaluation of current intervention strategies. As more information became available about the characteristics of the Delta and Omicron variants, many countries adapted their vaccination strategies to prioritise booster doses [9, 10], partly due to decreased vaccine effectiveness against new variants.

Mathematical modelling provides the tools to quantify how vaccine mechanisms and disease characteristics may affect the optimal impact of vaccine allocation. For simple epidemiological models such as the SIR or SEIR model, we can calculate the threshold vaccination needed to ensure *R*_0_ *<* 1 which ensures sustained transmission is not possible. While we can derive the threshold of vaccination to ensure *R*_0_ *<* 1 for more complex models, the relationship between various objectives such as total infections or deaths and *R*_0_ is no longer as simple as before. As such, calculating the number of vaccines needed to optimise a specific objective requires careful thought and potentially numerical methods. Realistically, we may instead compare a range of vaccination strategies to determine which results in the best outcome for a given objective, deeming this the optimal of the strategies considered [3, 7, 8, 11]. In general, the choice of public health objective will impact results, so we need to consider which metrics, such as total infections, total symptomatic infections or total deaths, are most appropriate to minimise when optimising a vaccine allocation strategy [12].

Two important areas related to pre-epidemic vaccination are exploring optimal vaccine allocation given disease characteristics and allocation objectives to prepare for potential pandemics, and designing optimal vaccine allocation strategies based on limited information from novel pandemics. To predict the optimal allocation strategy for potential pandemics, we want to know who to vaccinate and when to vaccinate. Vaccines are generally most effective when allocated early in the outbreak and rapidly, focusing primarily on vulnerable people and highly transmissible groups [3, 4, 6, 13, 14]. When a pandemic begins, we can apply the findings from this literature to predict effective strategies based on what information is available. For both influenza and COVID-19, optimal vaccine strategies depend on how transmissibility, severity and vaccine effectiveness vary by age [4, 11, 7, 15, 16, 8]. While these analyses focus on determining the optimal vaccine allocation strategy given pathogen characteristics, they do not present a generalised understanding of difference between optimal and suboptimal strategies. Are there certain situations where vaccine coverage should be prioritised, irrespective of who gets vaccinated? Or are there situations where we should ensure every available vaccine is allocated optimally?

In this paper we develop an approach to answer these vaccine allocation questions. We develop deterministic models and consider the effects of four separate vaccine mechanisms on a population comprised of two sub-groups with varying characteristics. We then determine the optimal allocation strategies for this population for a variety of allocation objectives. Through comparing the outcomes of optimal and suboptimal allocation strategies, we investigate how disease characteristics (such as *R*_0_ and disease severity) and vaccine characteristics (such as vaccine effectiveness and coverage) impact the differences in outcomes for various strategies. This analysis explores how the difference in outcomes from prioritising better vaccine resources or prioritising targeted allocation changes depending on the scenario considered.

## 2. Method

We define a modified SEIR model which includes two population groups, pre-epidemic vaccination, symptomatic and asymptomatic infection, and death due to disease. We consider vaccine mechanisms that prevent either transmission, symptomatic infection or severe disease, and consider scenarios where people are vaccinated before an epidemic starts. Through numerical simulation of our model, we aim to minimise either total infections, symptomatic infections or deaths and explore how disease and vaccine characteristics affect the outcome of a variety of vaccine allocation strategies.

### 2.1. Model assumptions and definition

We define our model from the following disease progression for each population group:

- Every person is initially susceptible, and are split into *unvaccinated* and *vaccinated* groups based on their pre-epidemic vaccination status.
- If infected, a susceptible person becomes *exposed*, where they are infected but not yet infectious.
- An exposed person will become infectious, developing either *symptomatic* or *asymptomatic disease*.
- A person with symptomatic disease can either *recover* from disease, or *die* due to infection.
- A person with asymptomatic disease will *recover* from disease.

Our model is described by the compartment diagram in Figure 1. We consider compartments

**Figure 1:**
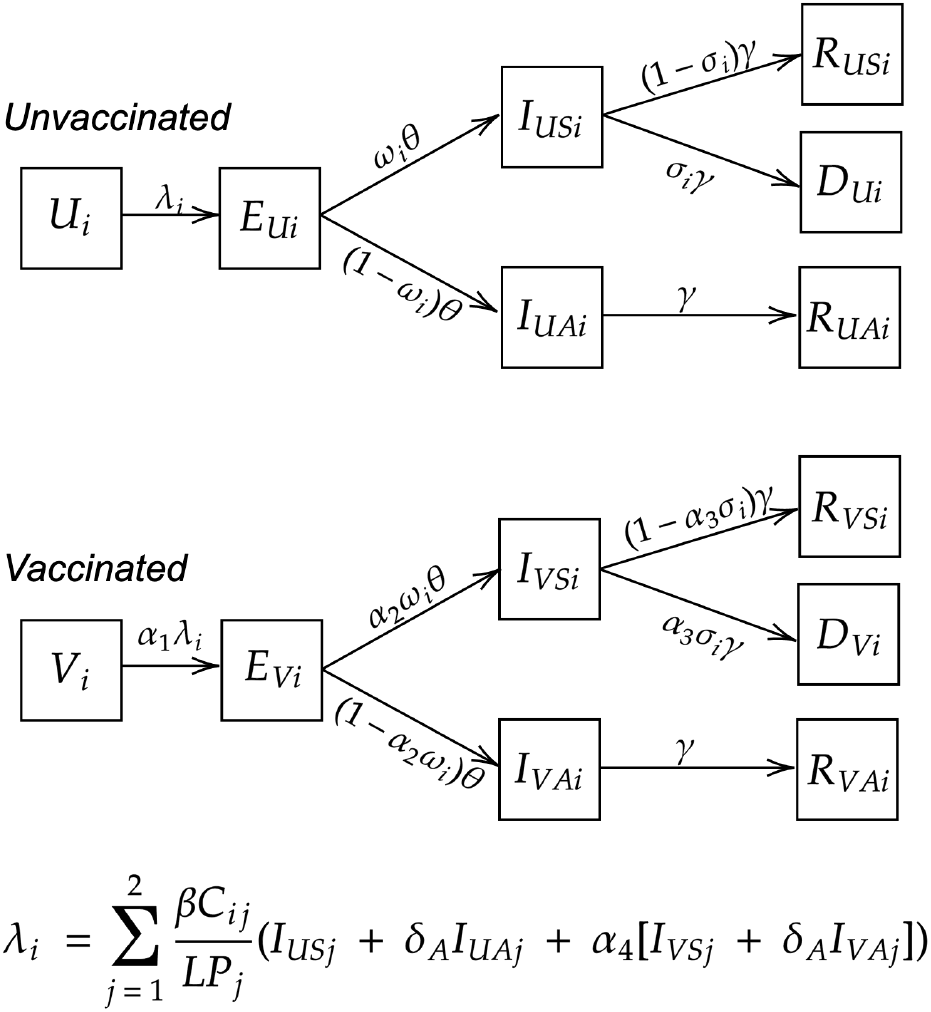
Compartmental diagram for our extended SEIR model. Initially people are either susceptible (U_i_) or vaccinated (V_i_) where i ∈ {1, 2} denotes population group. Both classes are able to become exposed (E_ji_) at rate and then infected (I_jki_) where j ∈ {U, V } denotes whether they were initially unvaccinated or vaccinated and k ∈ {S, A} denotes if they experience symptomatic or asymptomatic disease. From here, symptomatic people or asymptomatic can recover from disease (R_jki_) or symptomatic people can also die due to disease (D_ji_). As defined in the standard SEIR model, θ denotes the rate at which people transition from exposed to infected and γ denotes the rate at which people are removed from infected compartments. In this model, ω_i_ denotes the probability of developing symptomatic infection and σ_i_ denotes the probabiity of dying due to disease. We define vaccine effectiveness for each mechanism as 1 − α_m_ for m ∈ {1, 2, 3, 4}. We consider mechanisms: 1) reducing susceptibility, 2) reducing probability of developing symptomatic infection, 3) reducing probability of dying due to disease, and 4) reducing infectivity. In the force of infection λ_i_, βC_ij_ denotes the transmission rate between a susceptible in Group i and an infected in Group j. 1 − δ_A_ denotes the reduction in infectivity due to asymptomatic infection and LP_j_ denotes the number of people alive in Group j.

*X*_*jki*_, where:

- *X* ∈ {*U, V, E, I, R, D*}: denotes compartment types; *Unvaccinated, Vaccinated, Exposed, Infected, Recovered* and *Dead*.
- *j* ∈ {*U, V* }: denotes *Unvaccinated* or *Vaccinated* disease progression,
- *k* ∈ {*S, A*}: denotes *Symptomatic* or *Asymptomatic* infection, and
- *i* ∈ {1, 2}: denotes subpopulation group, denoted Group 1 and Group 2.

For example, the compartment *I*_*US*1_ is comprised of people in Group 1 who were initially unvaccinated and have developed symptomatic infection.

To illustrate the impact of vaccination on subpopulation groups with varying characteristics, we define Group 1 as twice as susceptible to infection the Group 2 (unless otherwise stated). We define the contact matrix *C* to characterise the interactions between susceptible and infectious individuals in each group:

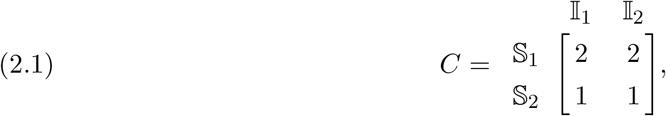

where 𝕊 = {*U*_*i*_, *V*_*i*_} denotes the susceptible compartments and 𝕀 = {*I*_*USi*_, *I*_*UAi*_, *I*_*V Si*_ and *I*_*V Ai*_} denotes the infected compartments. That is, we can define the force of infection *λ*_*i*_(*t*) for each subpopulation group *i* by:

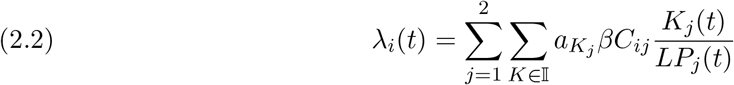

where *β* is the transmission rate, *a*_*Kj*_ scales *β* to suit the characteristics of compartment *K*_*j*_, and *LP*_*j*_(*t*) is the number of people alive in Group *j* at time *t*.

We choose to vary our transmission parameter *β* through varying the reproductive number, *R*_0_. We calculate *R*_0_ numerically for our model by finding the largest eigenvalue of the next generation matrix [17]. Using this calculation, we calibrate *β* for a given *R*_0_ value.

In this model, we assume frequency dependent transmission i.e. the transmission rate depends on the proportion of infecteds in the population as the contact rate is independent of population size. This is preferred for modelling human populations over density dependent transmission as we determine our contacts by social constraints, which do not necessarily scale with population size [5]. We further assume disease progresses on a fast enough timescale that we can ignore births and natural deaths and that the population is closed. Therefore, the total population for each group *P*_*i*∈{1,2}_ is constant, but this population also includes people who have died due to disease. To implement frequency dependent transmission, we define a new population size *LP*_*i*∈{1,2}_(*t*) which denotes the size of the *living* population for each group. This living population will change over time as people die due to disease. As people die and no longer have contact with living people (we assume), for the same number of infected people, the proportion of infected contacts will increase.

To investigate the effects of a range of vaccine mechanisms, we consider four vaccine mechanisms [16]:

1. Reduction in susceptibility,
2. Reduction in probability of developing symptomatic infection,
3. Reduction in probability of dying due to disease, and
4. Reduction in infectivity.

We define the effectiveness of each vaccine as 1 − *α*_*m*∈{1,2,3,4}_ ∈ [0, 1], where *m* denotes the vaccine mechanism, as numbered above. An 100% effective vaccine (1 − *α*_*m*_ = 1) corresponds to a perfect vaccine, for example if *α*_1_ = 0, vaccinated individuals are unable to be infected. Conversely, a 0% effective vaccine (1 − *α*_*m*_ = 0) corresponds to the vaccine having no effect. When vaccine effectiveness is not varied, we will assume each vaccine has a baseline values of *α*_*m*_ = 0.75. As we only consider pre-epidemic vaccination, vaccine allocation appears in the initial conditions for the vaccinated compartments for Groups 1 and 2 (*V*_1_ and *V*_2_). We assume those with asymptomatic infection are less infectious than those with symptomatic infection (this reduction in infectivity is given by the quantity 1 − *δ*_*A*_ = 0.5).

From the assumptions stated above, we derive the ODEs to describe our model. In addition to the standard SEIR model parameters, *α*_*m*_ denotes the vaccine effectiveness, *ω*_*i*_ denotes the proportion of exposed individuals that become symptomatically infected, *σ*_*i*_ denotes the proportion of symptomatic individuals who die due to disease, and 1−*δ*_*A*_ denotes the reduction in infectivity for asymptomatic infecteds, all for a population group *i*. Furthermore, *λ*_*i*_ denotes the force of infection, as defined by Eq. (2.2), *C*_*ij*_ denotes the elements of the contact matrix, as defined by Eq. (2.1), *P*_*j*_ denotes the size of Group *j* at the beginning of the epidemic, and *LP*_*j*_(*t*) denotes the size of the living population of Group *j*. Our model is hence by the following equations for each population group *i*:

#### Unvaccinated

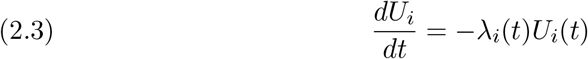

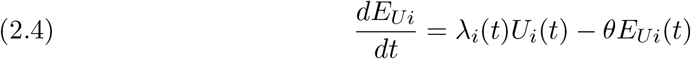

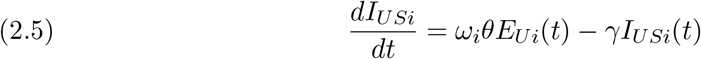

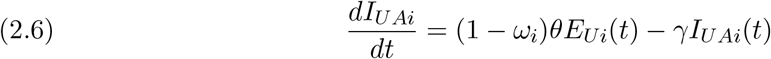

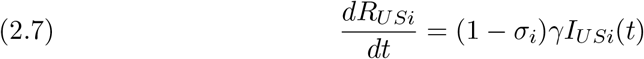

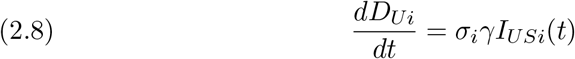

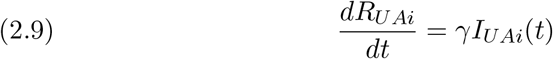

#### Vaccinated

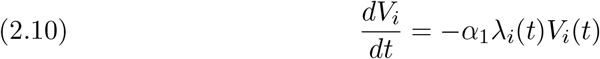

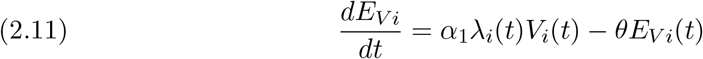

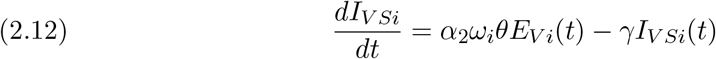

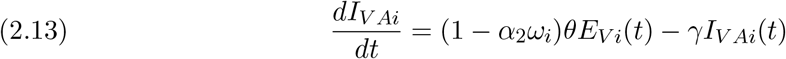

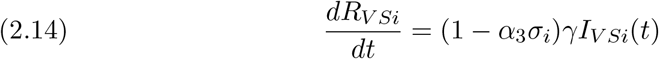

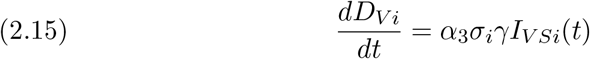

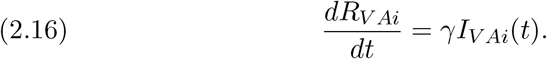

#### Force of infection

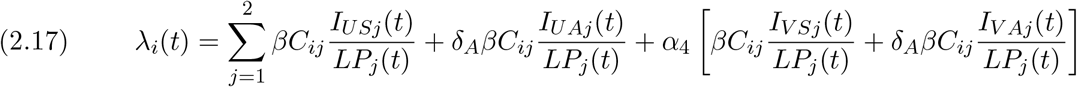

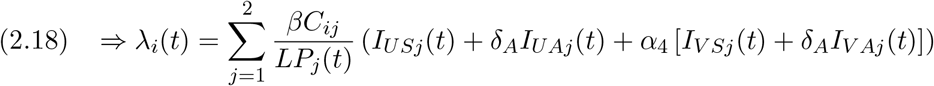

Adding Eqs. (2.3)–(2.16) we find:

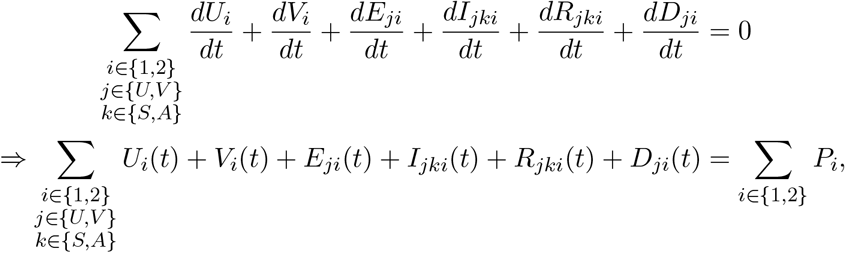

where we define *P*_1_ and *P*_2_ as the population sizes of Groups 1 and 2 at the beginning of the epidemic. For our analysis we assume *P*_1_ = *P*_2_. As the epidemic progresses and people die due to disease, we define the living population size *LP*_*i*∈{1,2}_(*t*) as:

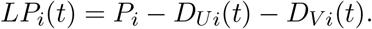

Given both Groups 1 and 2 start with *I*_0_ total infections each, and *v*_1_ and *v*_2_ vaccinations respectively, Eqns (2.3)–(2.16) have the following initial conditions:

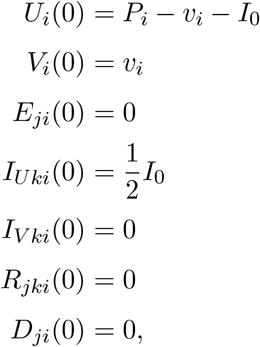

for *i* ∈ {1, 2}, *j* ∈ {*U, V* } and *k* ∈ {*S, A*}. That is, we assume there are initially 2*I*_0_ infected people with *I*_0_ infected people in each population group. For each population group all initially infected people are unvaccinated with half symptomatic and the other half asymptomatic. In our analysis we assume *I*_0_ = 50, that is there are 100 people initially infected across the whole population. Complete modelling details are given in the supplementary material.

### 2.2. Optimising Vaccine Allocation

To find the optimal vaccine allocation strategy, we compute the number of vaccines allocated to each subpopulation group to minimise a given objective function. Given we consider a finite number of vaccines (*v*_*max*_) and two subpopulation groups, the problem reduces to finding the number of vaccines allocated to Group 1 (*v*_*opt*_) that minimise the objective function:

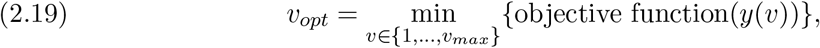

where *y* is the output from the model Eqs. (2.3)–(2.16) with initial conditions *V*_1_(0) = *v* and *V*_2_(0) = *v*_*max*_ − *v*.

We consider three objective functions:

1. Total infections:

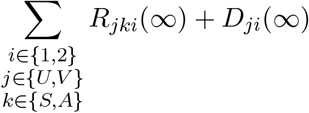
2. Total symptomatic infections:

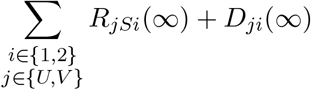
3. Total deaths:

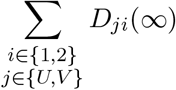

where 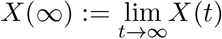 denotes the size of compartment *X* at the end of the epidemic.

We aim to compare optimal and suboptimal strategies as we vary disease and vaccine characteristics. To calculate the optimal strategy for any set of parameter values, we use the MATLAB function `fmincon` to find the proportion of vaccines allocated to each group that minimises the objective (total infections, total symptomatic infections, or total deaths). We found the optimal strategies for our model prioritised vaccinated one group over another. Using this optimal strategy, we compute sub-optimal strategies by varying the proportion of vaccines allocating to each group. In particular, we define a “poor strategy” as prioritizing vaccination for the opposite group to the optimal strategy. We also define an “uniformed strategy” as vaccinating each group equally.

When varying parameters in our model, we vary some continuously, and choose discrete examples for others. For continuously varying parameters, we compare the given objective for the optimal strategy, uninformed strategy and poor strategy. For parameters where we have chosen specific discrete examples, we compare a variety of strategies, chosen by varying the proportion of vaccines allocated to each group.

The results presented were generated using MATLAB 9.10, and our model was simulated using `ode45`.

## 3. Results

We present our results to investigate the following questions:

1. How do disease characteristics lead to variation in the differences in outcomes between optimal and suboptimal vaccination strategies?
2. How does the difference in outcome between optimal and suboptimal strategies depend on vaccine effectiveness and coverage?

When examining our results, it is useful to think about the direct and indirect effects of various parameters or vaccine mechanisms. We use “direct effects” to mean mechanisms that impact only vaccinated individuals, for example if the vaccine reduces the probability of hospitalisation. Conversely, we use “indirect effects” to refer to mechanisms that impact both vaccinated and unvaccinated individuals due to the vaccine’s impact on disease transmission, for example if vaccination results in reduced contagiousness.

The figures in Sections 3.1 and 3.2 show different types of parameters, either varied continuously or discretely. For certain parameters, such as *R*_0_ or *α*_*i*_, we can consider continuous variation when comparing the outcomes of vaccination strategies. These figures (Figures 2a, 3a and 3c), show straight and dotted lines to represent how the optimal, poor, and uniformed strategies perform as we vary our considered parameter. However, for some parameters, such as *σ*_*i*_ and vaccine coverage, we instead vary the parameters discretely. In this case, we choose discrete parameter values and consider the outcomes of a range of vaccination strategies (defined by assigning vaccines to each group in varying proportions) given these parameters. In these figures (Figures 2b, 3b and 3c) we use circles to represent the outcome of a single vaccination strategy. The range of these circles for each discrete parameter value gives us insight into the difference between the best and worst strategies for each scenario.

**Figure 2:**
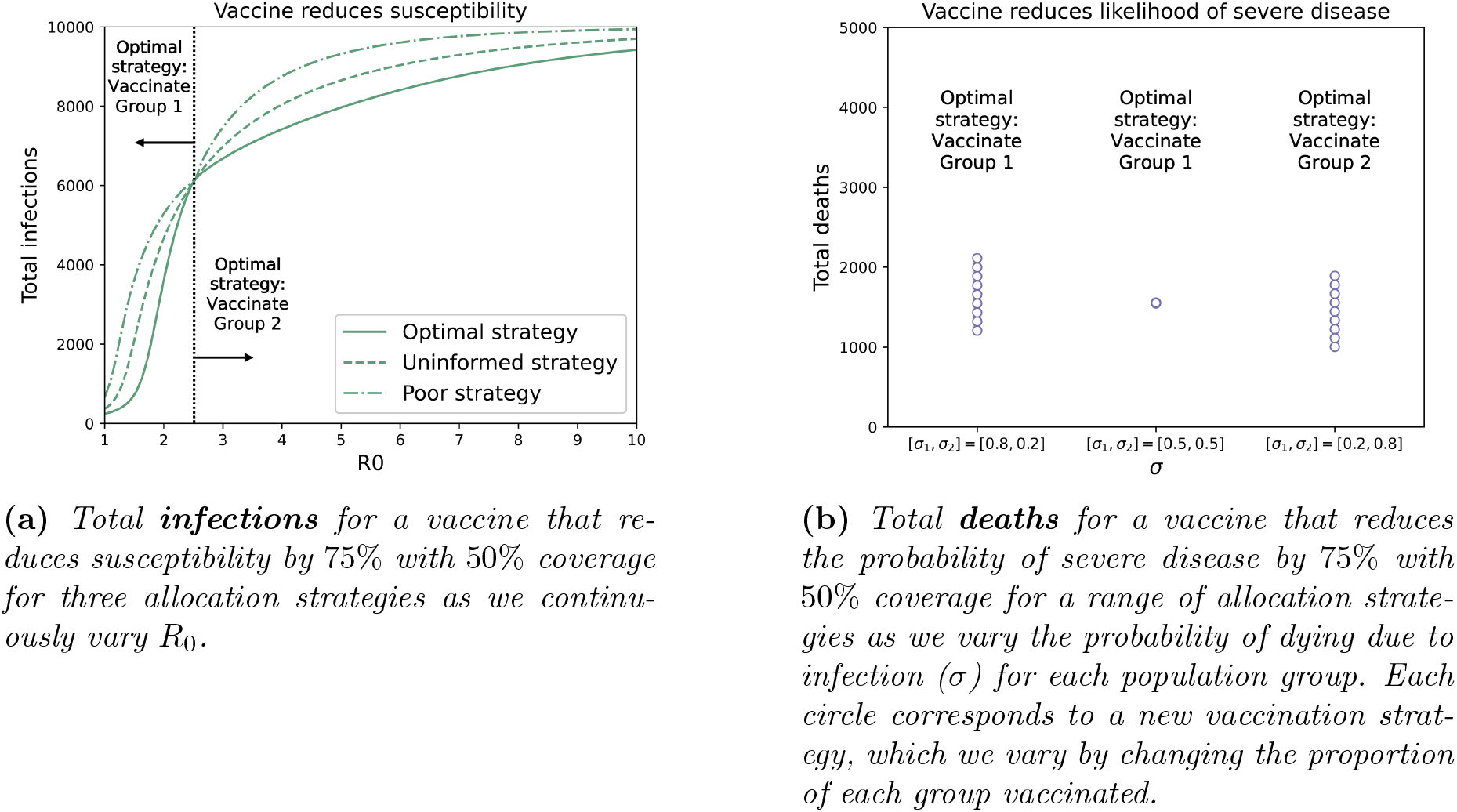
Comparing the differences between strategies for varying vaccine mechanisms and allocation objectives as we vary R_0_ and the probability of dying due to infection σ. Note the different allocation objectives for each figure.

In the following sections we provide a subset of modelling results, deliberately chosen to explore interesting aspects of our modelling. However, we also provide the complete set of model results in the supplementary material. The figures in section 2 of the supplementary material show how the outcomes of various vaccine strategies differ for all combinations of vaccine mechanism, allocation objective and considered parameter values.

### 3.1. Varying disease characteristics

Here we present results that consider how the difference between optimal and suboptimal strategies is dependent on disease characteristics for various vaccine mechanisms and allocation objectives. We consider vaccination strategies for a 75% effective vaccine with 50% vaccine coverage. These values have been chosen deliberately to demonstrate the effects of varying disease characteristics and are broadly reasonable in terms of typical vaccine effectiveness measures for respiratory diseases [18, 19, 20]. As we assume Group 1 and Group 2 are the same size, 50% vaccine coverage provides interesting results as it gives us the potential to vaccinate the entirety of one population group. Unless otherwise stated, we assume a total population of size of *N* = 10000, with 5000 people in each group (*P*_1_ = *P*_2_ = 5000).

The plots in Figure 2 show the effects of varying a parameter that affects transmission (reproduction number: *R*_0_) and another that affects infection characteristics (probability of dying due to infection: *σ*) on the difference between optimal and suboptimal allocation strategies. Note that Figures 2a and 2b have different allocation objectives as we are looking to compare how their respective objectives change between vaccination strategies as we vary parameter values.

We consider the effect of varying *R*_0_ on the difference in outcomes between strategies as shown in Figure 2a. There are two regions in this figure defined by a critical value 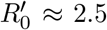 low 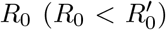 and high 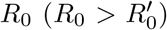. For low *R*_0_, there are few secondary infections (due to vaccination), and so the total infections is dominated by the number of initially infected people (*I*_0_). In our model, *I*_0_ = 20, and so the curves in Figure 2a are initially convex, which is not seen in typical final size curves due to the assumption of very small *I*_0_ (*I*_0_ ≈ 1% of the total population). For high *R*_0_ we recover the typical final size curve.

Qualitatively, we see a difference in outcome for all strategies as we vary *R*_0_. While the difference in outcome for high and low *R*_0_ depends on the strategy we choose, for all strategies total infections are monotonically increasing with *R*_0_. The importance of choosing the optimal strategy also varies with *R*_0_, with the largest differences between the optimal and poor strategies being when *R*_0_ is between 1 and 2. At 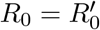, the outcomes of the optimal, uninformed and poor strategy overlap as the optimal strategy changes from prioritising vaccination for Group 1 to prioritising vaccination for Group 2. As *R*_0_ approaches the critical value of 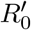, more people in Group 1 will be infected despite vaccination, closing the gap between the total infections from optimal and suboptimal strategies (more details can be found in the supplementary material). As *R*_0_ increases above 5, the outcome for all strategies gets consistently worse and the difference between strategies decreases.

In contrast, we consider the effect of varying the probability of dying due to infection (*σ*) for each population group on the difference in outcomes between strategies, shown in Figure 2b. We consider three scenarios *σ*_*i*_: *σ*_1_ *> σ*_2_, *σ*_1_ = *σ*_2_, and *σ*_1_ *< σ*_2_. The biggest difference in outcomes of strategies happens when *σ*_1_ ≠ *σ*_2_. When *σ*_1_ = *σ*_2_, the only factor influencing the difference in deaths between population groups is their relative susceptibility (as defined previously). Hence, while there is a difference in the number of people infected in each population group, vaccination will reduce the probability of dying due to disease to the same value for each group. However, when *σ*_1_ ≠ *σ*_2_, one group is more likely to die due to disease. Therefore, vaccinating the group more likely to die due to disease will reduce more deaths than vaccinating the other group, widening the gap between the outcomes of alternative strategies. For parameters with only direct effects, such as *σ*_*i*_, vaccination only affects vaccinated people. The difference between outcomes of allocation strategies is therefore dependent on how the parameters for each group compare to one another, and is not influence by the non-linear infection dynamics.

The two examples presented in Figure 2 demonstrate how direct and indirect affects of vaccination impact the differences in outcomes between optimal and suboptimal strategies. For parameters affecting transmission (e.g. *R*_0_) the difference between outcomes of strategies is dependent on the value of the parameter itself. This is due to its affect on transmission for both population groups. However, for parameters affecting individual infection (e.g. *σ*_*i*_) the difference between outcomes of strategies is dependent on the relative parameter values between each population group. The biggest differences between outcomes of strategies will occur where there is large disparity in the parameter between groups, and the smallest difference when both groups are equal.

### 3.2. Varying vaccine characteristics

We now consider an example population to demonstrate how vaccine effectiveness and coverage affect the differences in outcomes between allocation strategies. We define Group 1 to be more susceptible, more likely to develop symptomatic disease and more likely to die due to disease, and Group 2 to be less susceptible, less likely to develop symptomatic disease and less likely to die due to disease.

Figure 3 compares the differences between allocation strategies for various vaccine mechanisms and allocation objectives as we vary vaccine effectiveness and coverage. Figures 3a and 3b show the total infections for vaccines that affect transmission as vaccine effectiveness and coverage are varied. For the considered parameters, the optimal strategy changes from prioritising vaccination for Group 2 to prioritising vaccination for Group 1 at 63% effectiveness (3a). For both vaccine effectiveness and coverage, as we saw in Figure 2a, as we increase our vaccine parameter we reduce our objective for all allocation strategies. As the vaccine tends towards becoming 100% effective, or there is high coverage (80%), the optimal strategy produces a much better result than other strategies. However, for lower effectiveness and coverage, increasing either parameter decreases our objective for all strategies. This demonstrates that, under the assumptions of our model, securing better vaccines or more vaccines will work to decrease our objective, even under a suboptimal strategy.

**Figure 3:**
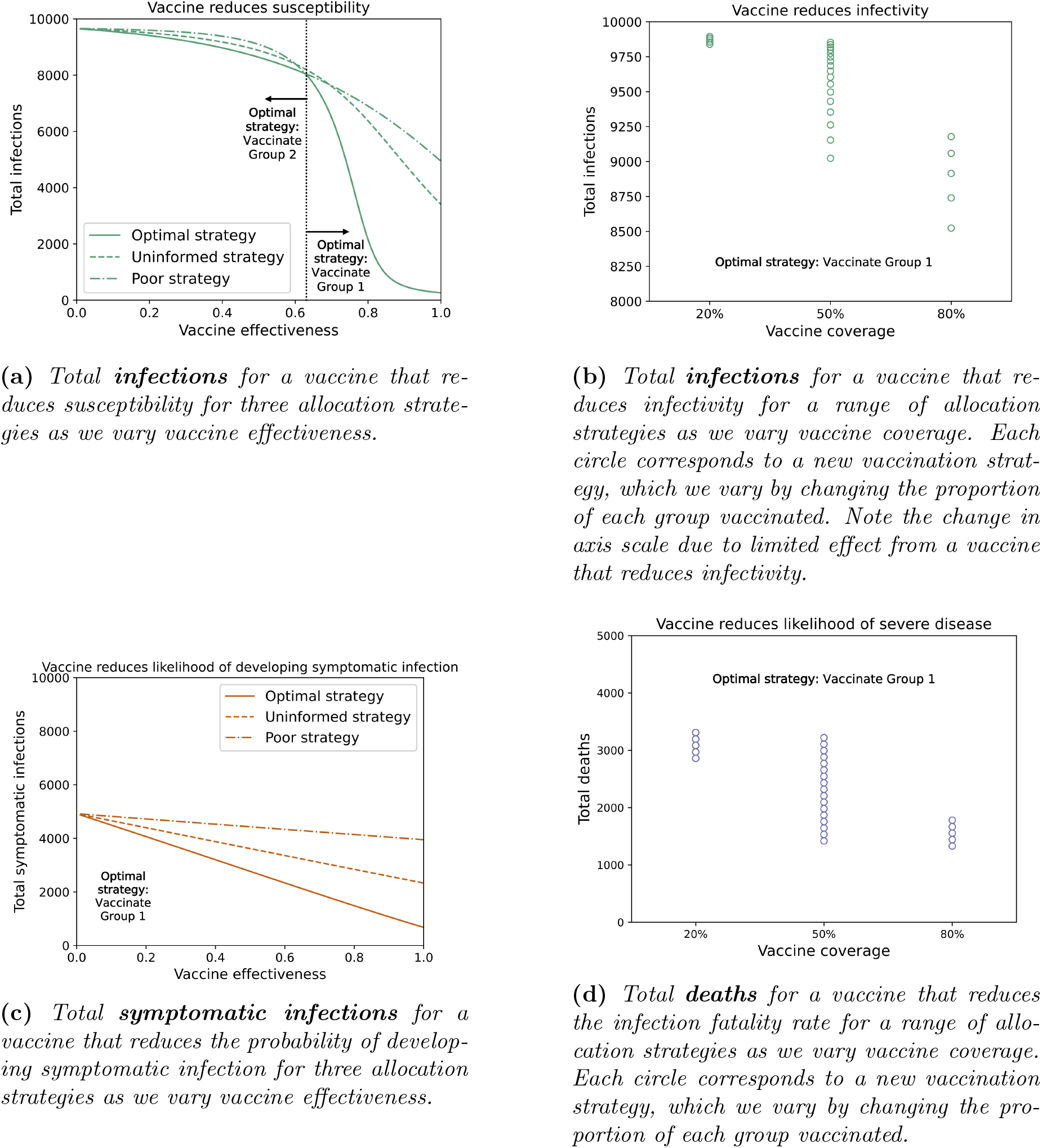
Comparing the differences between strategies for varying vaccine mechanisms and allocation objectives as we vary vaccine effectiveness and coverage. Each figure gives an example of how the difference between the outcomes of optimal and suboptimal strategies change between vaccine mechanisms that impact transmission, and mechanisms that affect individual infection. Note the different allocation objectives for each figure, and the reduced scale for Figure 3b.

Figures 3c and 3d show how total symptomatic infections and total deaths for vaccines that affect individual infection vary and we change vaccine effectiveness and coverage. In Figure 3d the difference between optimal and suboptimal strategies is much smaller for 20% and 80% coverage than 50% as there are fewer ways to allocate vaccines for 20% and 80% coverage than for 50% coverage. As vaccine effectiveness and coverage increase, there is a large reduction in outcome for optimal (and close to optimal) strategies, but a much smaller improvement for poor strategies. As vaccination only impacts infected people, vaccinating the less vulnerable, who are already unlikely to experience symptomatic or severe disease, does little to reduce an objective focused on these infection characteristics. Unlike a vaccine affecting transmission, where increasing quality or quantity of vaccines works to decrease the objective for all strategies, here there is only a sizeable reduction for the best strategies. This indicates that strategy remains important even when effective vaccines and high coverage is available. Our model results demonstrate that allocating poorly with an effective vaccine or high coverage could result in a much worse outcome than allocating well with an ineffective vaccine or lower vaccine coverage.

For our model and the parameters we chose, the difference between optimal and suboptimal strategies varies as we change vaccine effectiveness and coverage based on both vaccine mechanism and allocation objective. If a vaccine mechanism affects transmission, this will impact the total number of infections, and therefore also the total symptomatic infections and total deaths resulting from these infections. Hence for any of these objectives, increasing vaccine effectiveness or coverage will result in a better outcome for all strategies. However, if a vaccine mechanism only results in direct effects, this will only impact objectives associated with the mechanism. For example, a vaccine that only reduces the probability of dying due to disease will have no impact on total infections or total symptomatic infections. When the vaccine mechanism does impact an objective, increasing vaccine effectiveness and coverage is only effective when an optimal, or close to optimal, strategy is employed.

## 4. Discussion

Our results demonstrate that the difference between outcomes of vaccine allocation strategies depends on both disease and vaccine characteristics. The difference in outcomes between strategies is dependent on whether a considered parameter directly or indirectly impacts the outcome. For the former the difference between strategies depends heavily on the parameter value itself (e.g. the value of *R*_0_), whereas for the latter the difference depends on the how a parameter compares between population groups (e.g. which group is at higher risk of severe disease). Furthermore, a vaccine with direct effects will only impact vaccinated people, and so in our model poor vaccine resources allocated well can result in a better outcome than good vaccine resources allocated poorly. Conversely, if vaccination indirectly effects the outcome, better vaccine resources will result in a better outcome for all strategies. These findings emphasise the importance of identifying the direct and indirect effects of vaccination when determining allocation strategy.

By investigating allocation strategies for individual vaccine mechanisms and objectives, we lay foundations for a model that considers multi-mechanism or multi-objective strategies. When designing vaccine strategies with little knowledge about the specifics of the vaccine, modelling often assumes individual mechanisms, or simply that a vaccine provides reduced susceptibility [7, 15, 16]. When vaccines eventually become available and are observed to have multiple characteristics – against susceptibility, infectivity, and/or disease for example – appropriate mechanisms are built into models and parameterised using effectiveness parameters derived from epidemiological/clinical data [21, 22, 20]. Our model incorporates multiple possible vaccine mechanisms, however we restrict our analysis to the impact of each separately.

Furthermore, we consider objectives that are not strictly independent of one another. Due to our model assumptions, reducing infections leads to a reduction in symptomatic infections which further leads to a reduction in deaths. However, by considering the impacts of vaccination on transmission or individual infection, we can determine the impact of various disease and vaccine characteristics on the outcome of independent objectives. To consider allocation strategies for multiple objectives would require development of a single objective through weighting individual objectives based on importance. The impact of vaccination on this overarching objective could then be thought of in terms of the impact of vaccination on each individual part, as described in our results.

Our model was designed to explore the importance of different vaccine mechanisms. However, it was not intended to, nor does it describe any one specific realistic scenario. In our model, we assumed SEIR dynamics and divided the population into two sub-groups with differing characteristics. To model a more realistic scenario, we would likely need to extend our model to capture a more complex population and transmission structure. In particular, we assumed constant vaccine effectiveness across our population, but age dependent vaccine effectiveness has been observed, for example for influenza vaccines [23]. When considered in the model, this may alter not only the optimal vaccination strategy, but also the difference between optimal and suboptimal strategies [3, 16, 24]. The simplicity of our model allows us to analyse the differences in outcomes between optimal and suboptimal strategies, and identify important considerations for applied studies. Of course, more information and a model incorporating observed heterogeneities would be required to produce results for any vaccination scenario against a particular disease.

By investigating how various factors impact the outcomes of vaccination strategies in our model, we explore the idea of prioritising vaccine allocation over ascertainment. When new scenarios arise, such as new vaccines available or new variants appearing, our results allows us to compare how allocation strategies differ in this new scenario compared to an old scenario. Is it still worth sourcing more vaccines with the current allocation strategy or will increased coverage only improve the outcome if we vaccinate more effectively? How great are the benefits from allocating vaccines optimally now that our vaccine is less effective against a new variant? Do suboptimal strategies perform similarly? Our modelling shows that investigating how individual disease and vaccine characteristics impact the difference in outcomes between vaccination strategies is essential when considering development of, or updates to, vaccine policies. Regular reassessment of vaccine allocation as information on an emerging disease becomes available is important, reinforcing the need to carefully consider allocation strategy when distributing vaccines.

## Data Availability

The MATLAB code used to generate all results in this manuscript is available online.

https://github.com/iabell/vaccine_allocation

## Code availability

The MATLAB code used to generate all results in this manuscript is available online https://github.com/iabell/vaccineallocation.

## Supplementary materials

Additional figures and model details can be found in the supplementary materials.

