## Supplementary Material for "Understanding the impact of disease and vaccine mechanisms on the importance of optimal vaccine allocation"

### S1 Model details

In this section we briefly restate our model and parameter definitions, and we list the default values for each parameter. This is to provide the full model in a single, succinct section. We then provide extra results in Section S2.

We consider the model as defined by Eqs. (2)–(16):

$$\textit{Unvaccinated:} \tag{1}$$

$$\frac{dU_i}{dt} = -\lambda_i(t)U_i(t) \tag{2}$$

$$\frac{dE_{Ui}}{dt} = \lambda_i(t)U_i(t) - \theta E_{Ui}(t) \tag{3}$$

$$\frac{dI_{USi}}{dt} = \omega_i \theta E_{Ui}(t) - \gamma I_{USi}(t) \tag{4}$$

$$\frac{dI_{UAi}}{dt} = (1-\omega_i)\theta E_{Ui}(t) - \gamma I_{UAi}(t) \tag{5}$$

$$\frac{dR_{USi}}{dt} = (1-\sigma_i)\gamma I_{USi}(t) \tag{6}$$

$$\frac{dD_{Ui}}{dt} = \sigma_i \gamma I_{USi}(t) \tag{7}$$

$$\frac{dR_{UAi}}{dt} = \gamma I_{SAi}(t) \tag{8}$$

$$\textit{Vaccinated:} \tag{9}$$

$$\frac{dV_i}{dt} = -\alpha_1 \lambda_i(t)V_i(t) \tag{10}$$

$$\frac{dE_{Vi}}{dt} = \alpha_1 \lambda_i(t)V_i(t) - \theta E_{Vi}(t) \tag{11}$$

$$\frac{dI_{VSi}}{dt} = \alpha_2 \omega_i \theta E_{Vi}(t) - \gamma I_{VSi}(t) \tag{12}$$

$$\frac{dI_{VAi}}{dt} = (1-\alpha_2\omega_i)\theta E_{Vi}(t) - \gamma I_{VAi}(t) \tag{13}$$

$$\frac{dR_{VSi}}{dt} = (1-\alpha_3\sigma_i)\gamma I_{VSi}(t) \tag{14}$$

$$\frac{dD_{Vi}}{dt} = \alpha_3 \sigma_i \gamma I_{VSi}(t) \tag{15}$$

$$\frac{dR_{VAi}}{dt} = \gamma I_{VAi}(t) \tag{16}$$

*Force of Infection:*

$$\lambda_i = \sum_{j=1}^{2} \beta C_{ij} \frac{I_{USj}(t)}{LP_j(t)} + \delta_A \beta C_{ij} \frac{I_{UAj}(t)}{LP_j(t)} + \alpha_4 \left[ \beta C_{ij} \frac{I_{VSj}(t)}{LP_j(t)} + \delta_A \beta C_{ij} \frac{I_{VAj}(t)}{LP_j(t)} \right], \tag{17}$$

where $LP_i(t)$ denotes the living population at time $t$. That is,

$$LP_i(t) = P_i - D_{Ui}(t) - D_{Vi}(t),$$

where $P_i$ is the total population of group $i$ at $t = 0$.

### S1.1 Parameter definitions

We firstly define the compartments ($X_{jki}$) included in our model:

- $X \in \{U, V, E, I, R, D\}$: unvaccinated, vaccinated, exposed, infected, recovered, dead
- $j \in \{U, V\}$: unvaccinated or vaccinated disease progression
- $k \in \{S, A\}$: symptomatic or asymptomatic infection
- $i \in \{1, 2\}$: population group (1: high risk of infection, 2: low risk of infection)

The following parameters are defined to be constant across the population groups:

- $1/\theta = 1$: average time spent exposed
- $1/\gamma = 1$: average time spent infected
- $R_0 = 2$ (unless otherwise stated): reproduction number
- $\delta_A = 0.5$ (unless otherwise stated): reduction in infectivity due to asymptomatic infection

The following parameters are defined as specific to each population group $i$:

- $\omega_i = 0.5$ (unless otherwise stated): proportion of infected people in group $i$ that experience symptomatic disease
- $\sigma_i = 0.5$ (unless otherwise stated): proportion of symptomatic infecteds in group $i$ that die due to disease
- $P_i = 5000$: initial population size for group $i$
- $[C_{ij}] = \begin{bmatrix} 2 & 2 \\ 1 & 1 \end{bmatrix}$ (unless otherwise stated): Contact matrix
- $I_{0i} = 50$: Initial infected people in group $i$ (half symptomatic, half asymptomatic)

Finally, the following parameters characterise vaccination in our model:

- Vaccine coverage = 5000 (unless otherwise stated): Number of vaccines available
- $1 - \alpha_i = 0.25$ (unless otherwise stated): effectiveness of vaccine $i$
- $1 - \alpha_1$: reduction in susceptibility from Vaccine 1

- $1 - \alpha_2$: reduction in proportion of infected cases that experience symptomatic infection from Vaccine 2
- $1 - \alpha_3$: reduction in proportion of symptomatic infecteds that die due to infection from Vaccine 3
- $1 - \alpha_4$: reduction in infectivity from Vaccine 4

#### S1.2 Implicit Assumptions

In our model formulation, we assume that asymptomatic infection is less infective than symptomatic infection. This reduction in infectivity due to asymptomatic infection is denoted by $1 - \delta_A$. For this work we assume $\delta_A > 0$.

Furthermore, we assume people experiencing asymptomatic infection do not experience severe disease. This is implicitly assumed in our model as asymptomatic infecteds can only transition to a recovered compartment, they cannot die due to disease.

### S2 Results

#### S2.1 Figure 2a details

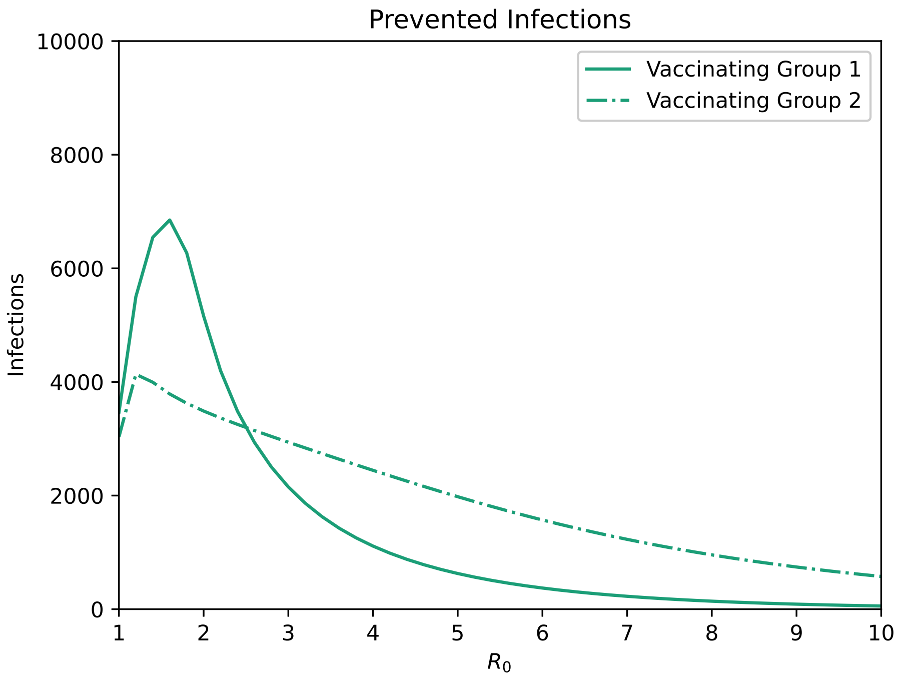


**Figure S1:** *Prevented infections from vaccinating either Group 1 or Group 2. The number of prevented infections is calculated by subtracting the total number of infections with vaccination from the total number of infections without vaccination.*

In this section we investigate the intersection point that appears between outcomes of vaccination strategies in Figure 2a. Consider Figure S1, which shows the prevented infections from vaccinating either Group 1 or Group 2. We see that before the intersection point ($R_0' \sim 2.5$), vaccinating Group 1 prevents the highest number of infections and is hence the optimal strategy. For $R_0 > R_0'$, vaccinating Group 2 becomes the optimal strategy, preventing more deaths than vaccinating Group 1. In Figure 2a, we assume Group 1 is more susceptible to disease than Group 2. As such, for $R_0 < R_0'$, there are few Group 2 infections to begin with, and so the optimal strategy is to vaccinate Group 1 to reduce their many infections. However, as $R_0$ increases, the number of people infected in Group 1 increases until almost everyone is infected, even if the entire group is vaccinated. We can see this in Figure S1, where as we increase $R_0$, we see diminishing returns from vaccinating Group 1, and we see more prevented infections from vaccinating Group 2. The reduced susceptibility of Group 2, compared to Group 1, means that it will take higher $R_0$ for the entirety of Group 2 to become infected, especially if vaccinated. Hence, we see more prevented infections from vaccinating Group 2 for $R_0 > R_0'$ than from vaccinating Group 1.

### S2.2 Extra plots

Here we show how the difference between strategies differs for every combination of vaccine mechanism and allocation objective as we vary specific parameters.

- Varying the contact matrix (Subsection S2.2.1)
- Varying $\omega_i$ (Subsection S2.2.2)
- Varying $\sigma_i$ (Subsection S2.2.3)
- Varying $\delta_A$ (Subsection S2.2.4)
- Varying $R_0$ (Subsection S2.2.5)
- Varying vaccine coverage (Subsection S2.2.6)
- Varying vaccine effectiveness (Subsection S2.2.7)

#### S2.2.1 Varying the contact matrix

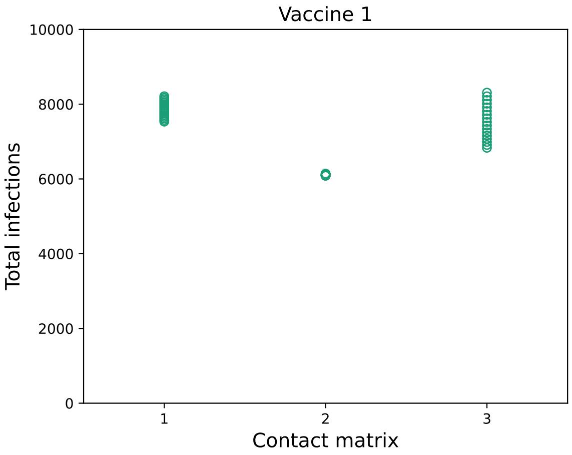


**(a)**

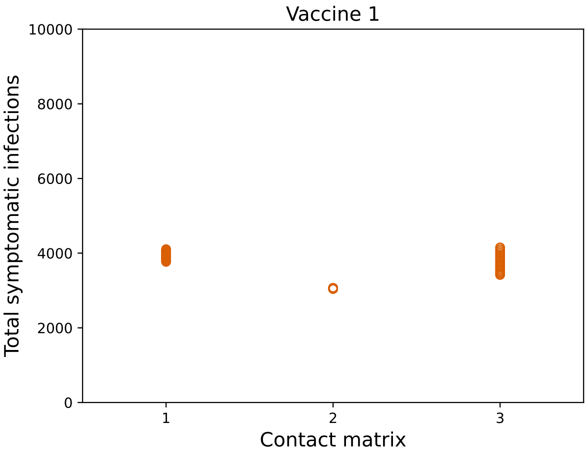


**(b)**

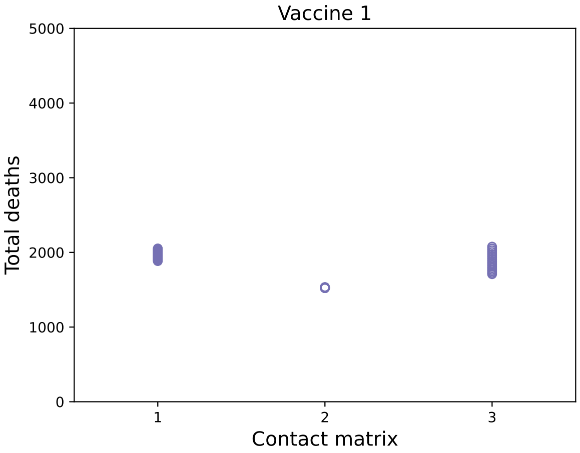


**(c)**

**Figure S2:** *Comparing total infections, symptomatic infections and deaths for Vaccine 1 as we vary contact matrices. The three contact matrices considered are (1)* $\begin{bmatrix} 4 & 2 \\ 2 & 1 \end{bmatrix}$, *(2)* $\begin{bmatrix} 2 & 2 \\ 1 & 1 \end{bmatrix}$, *and (3)* $\begin{bmatrix} 2 & 4 \\ 1 & 2 \end{bmatrix}$.

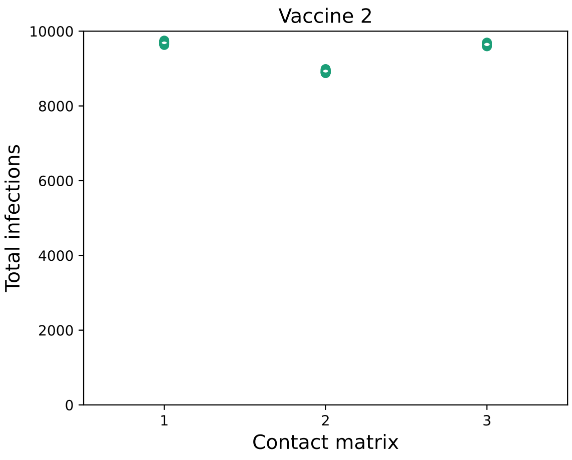


**(a)**

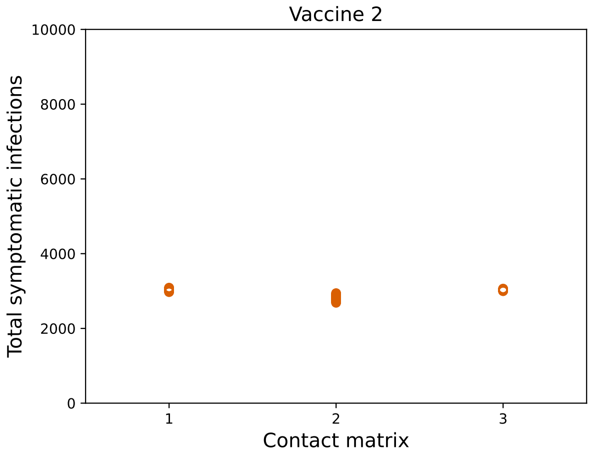


**(b)**

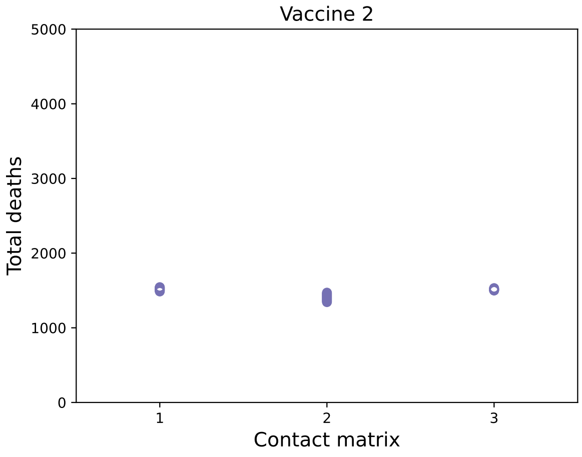


**(c)**

**Figure S3:** *Comparing total infections, symptomatic infections and deaths for Vaccine 2 as we vary contact matrices. The three contact matrices considered are (1)* $\begin{bmatrix} 4 & 2 \\ 2 & 1 \end{bmatrix}$, *(2)* $\begin{bmatrix} 2 & 2 \\ 1 & 1 \end{bmatrix}$, *and (3)* $\begin{bmatrix} 2 & 4 \\ 1 & 2 \end{bmatrix}$.

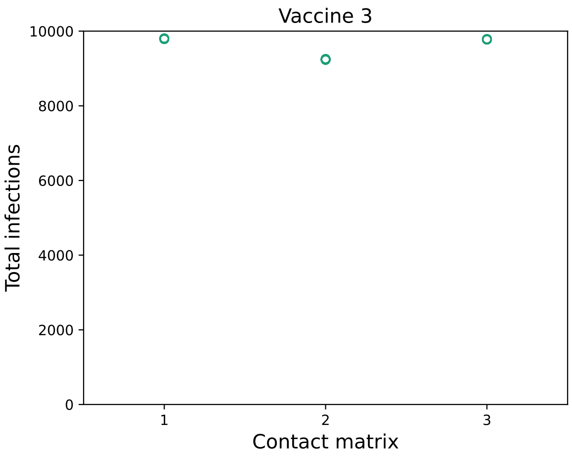


**(a)**

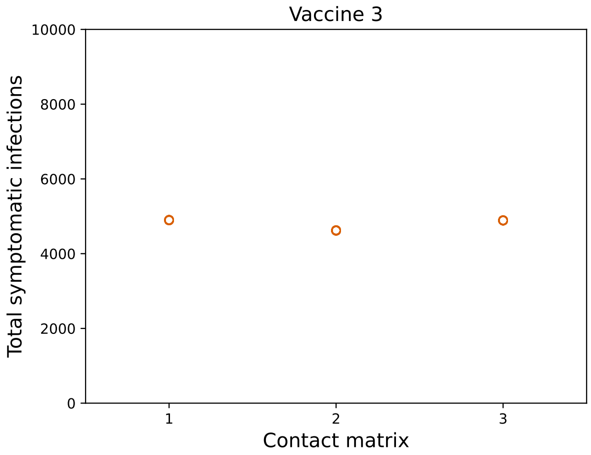


**(b)**

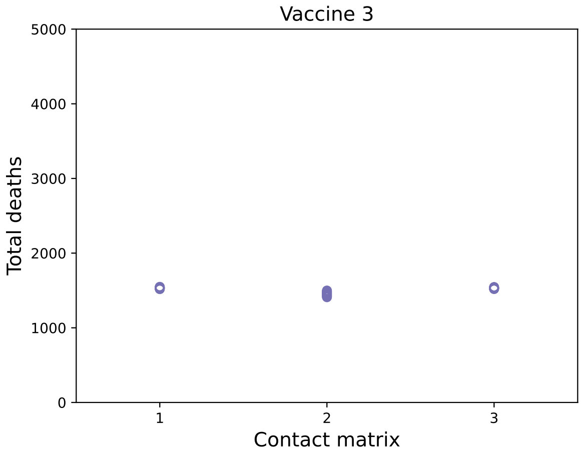


**(c)**

**Figure S4:** *Comparing total infections, symptomatic infections and deaths for Vaccine 3 as we vary contact matrices. The three contact matrices considered are (1)* $\begin{bmatrix} 4 & 2 \\ 2 & 1 \end{bmatrix}$, *(2)* $\begin{bmatrix} 2 & 2 \\ 1 & 1 \end{bmatrix}$, *and (3)* $\begin{bmatrix} 2 & 4 \\ 1 & 2 \end{bmatrix}$.

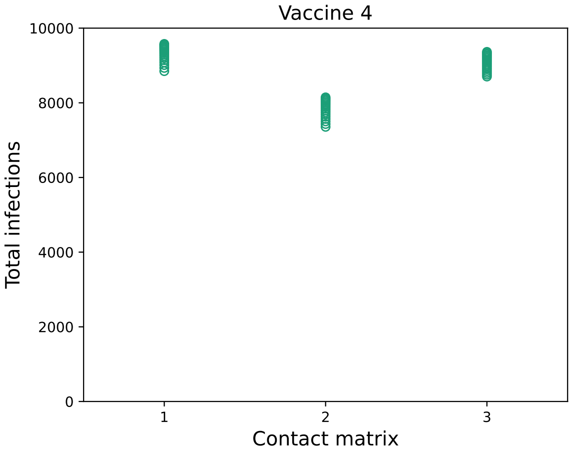


**(a)**

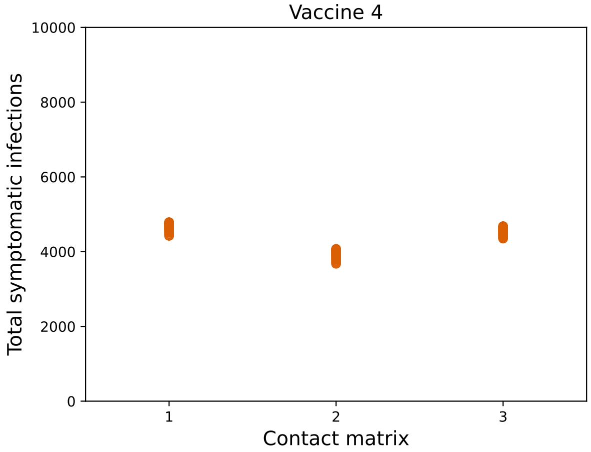


**(b)**

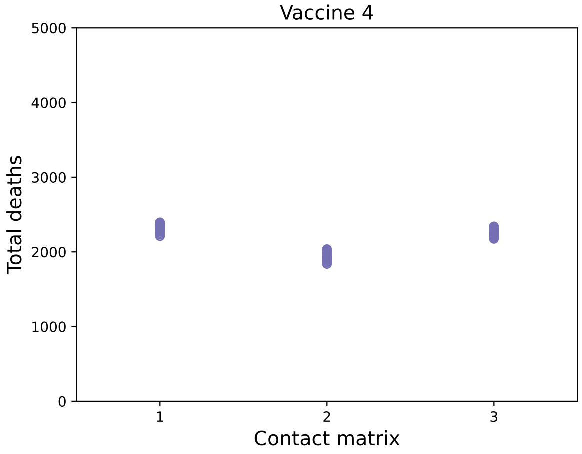


**(c)**

**Figure S5:** *Comparing total infections, symptomatic infections and deaths for Vaccine 4 as we vary contact matrices. The three contact matrices considered are (1)* $\begin{bmatrix} 4 & 2 \\ 2 & 1 \end{bmatrix}$, *(2)* $\begin{bmatrix} 2 & 2 \\ 1 & 1 \end{bmatrix}$, *and (3)* $\begin{bmatrix} 2 & 4 \\ 1 & 2 \end{bmatrix}$.

#### S2.2.2 Varying $\omega_i$

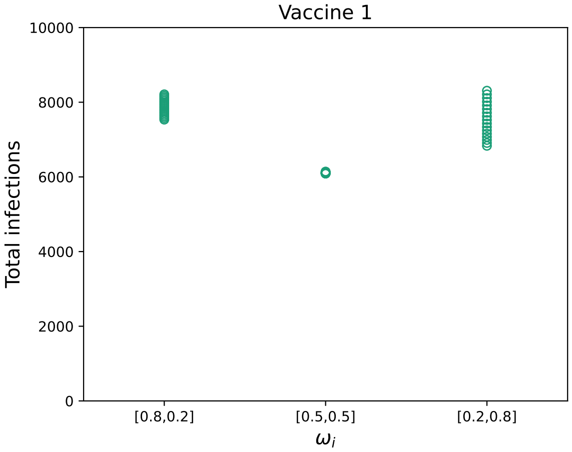


(a)

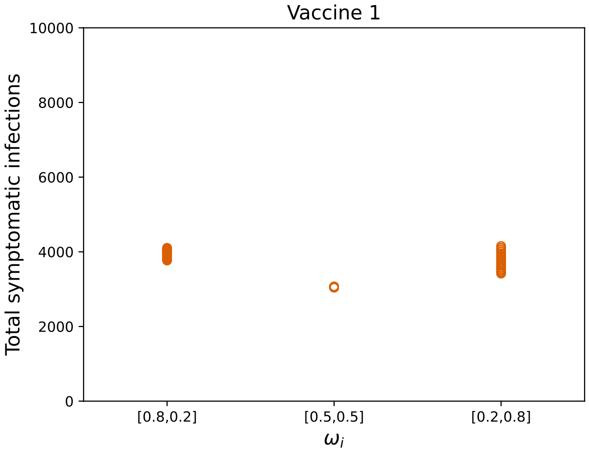


(b)

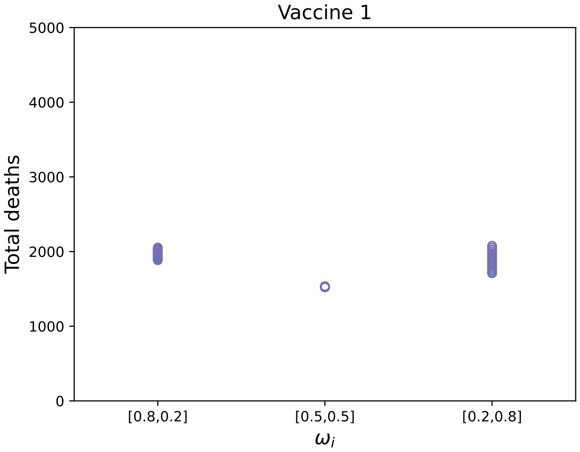


(c)

**Figure S6:** *Comparing total infections, symptomatic infections and deaths for Vaccine 1 as we vary* $\omega_1$ *and* $\omega_2$*.*

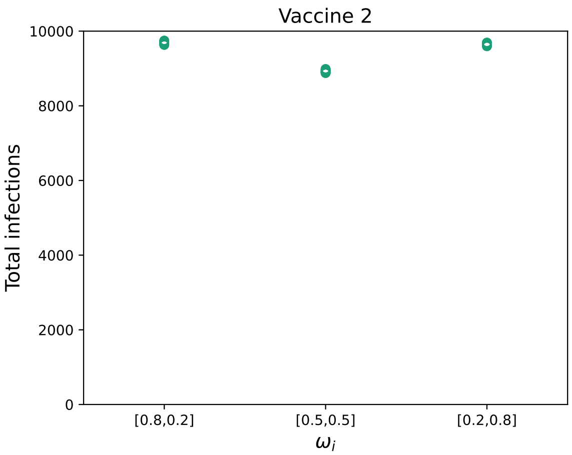


**(a)**

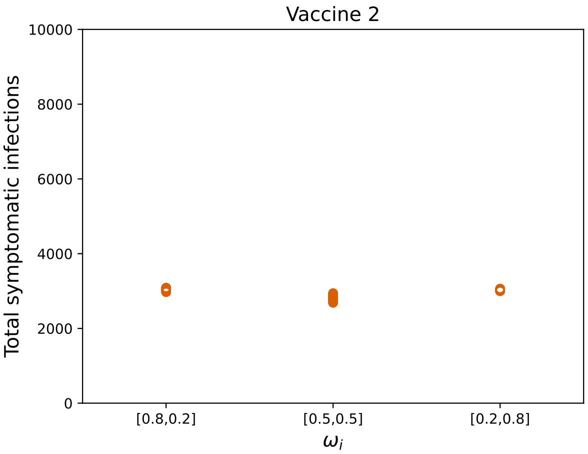


**(b)**

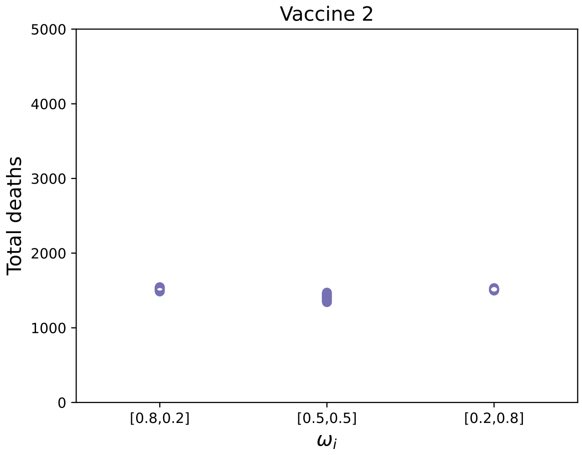


**(c)**

**Figure S7:** *Comparing total infections, symptomatic infections and deaths for Vaccine 2 as we vary $\omega_1$ and $\omega_2$.*

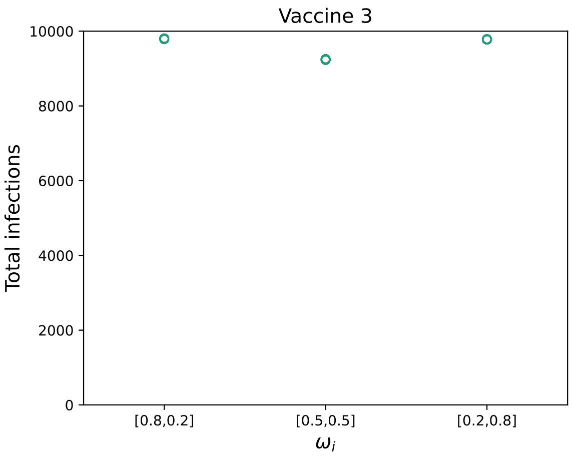


**(a)**

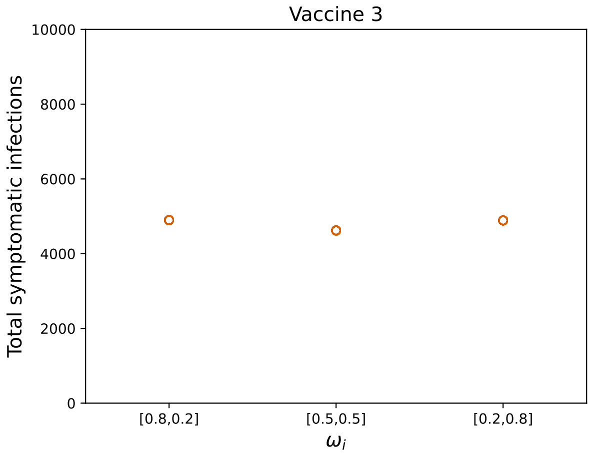


**(b)**

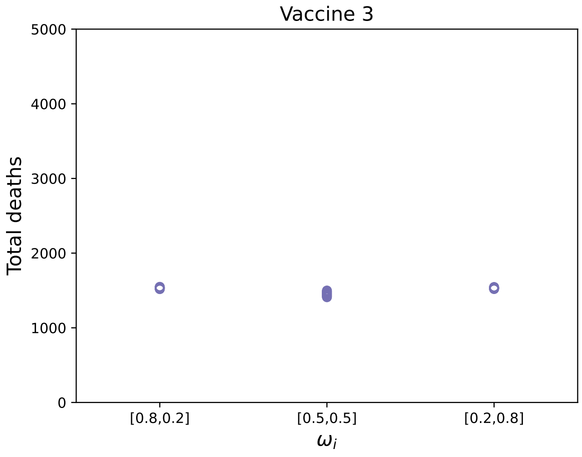


**(c)**

**Figure S8:** *Comparing total infections, symptomatic infections and deaths for Vaccine 3 as we vary* $\omega_1$ *and* $\omega_2$.

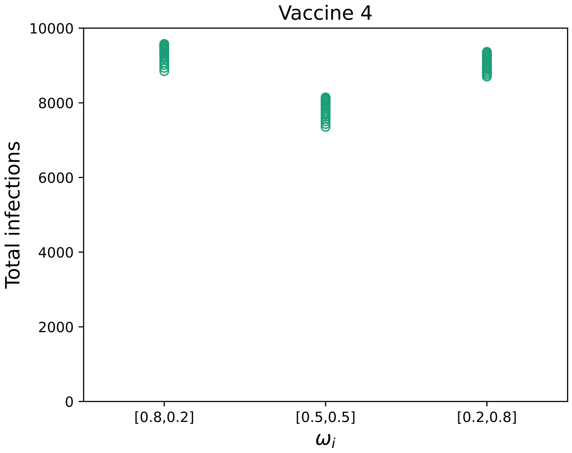


**(a)**

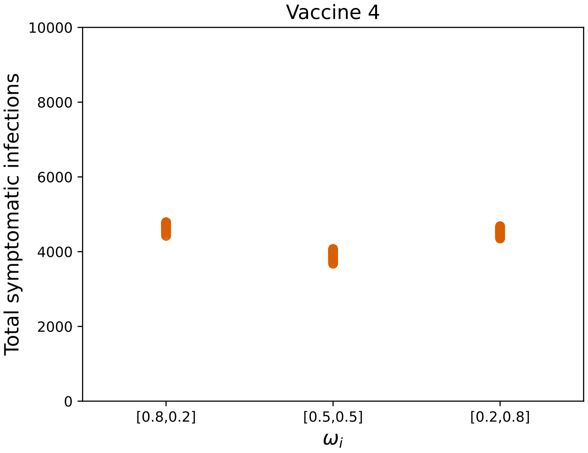


**(b)**

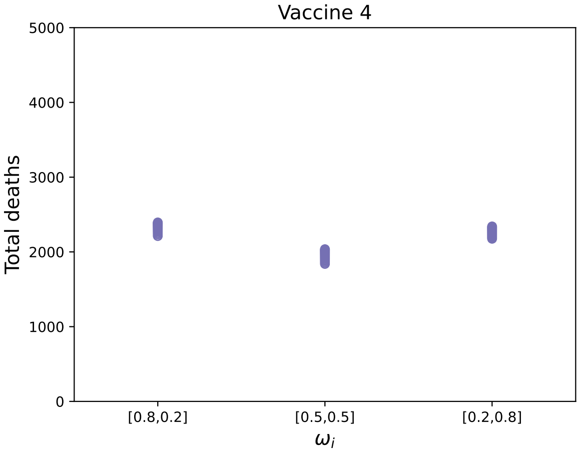


**(c)**

**Figure S9:** *Comparing total infections, symptomatic infections and deaths for Vaccine 4 as we vary* $\omega_1$ *and* $\omega_2$.

#### S2.2.3 Varying $\sigma_i$

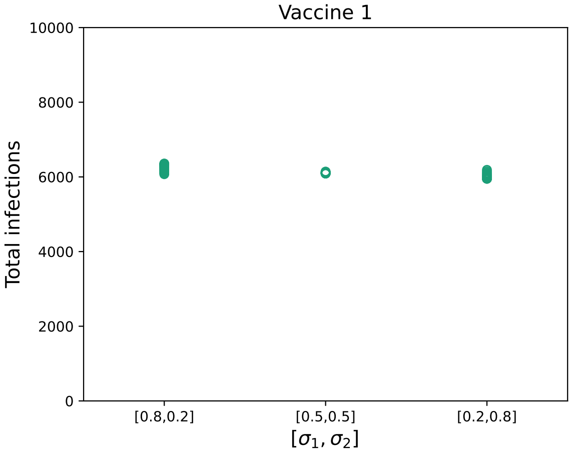


**(a)**

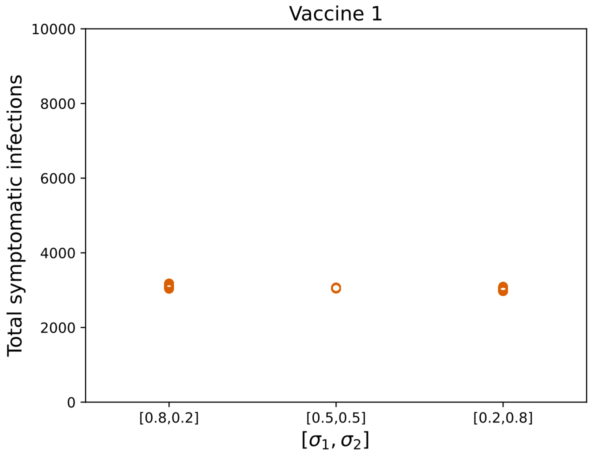


**(b)**

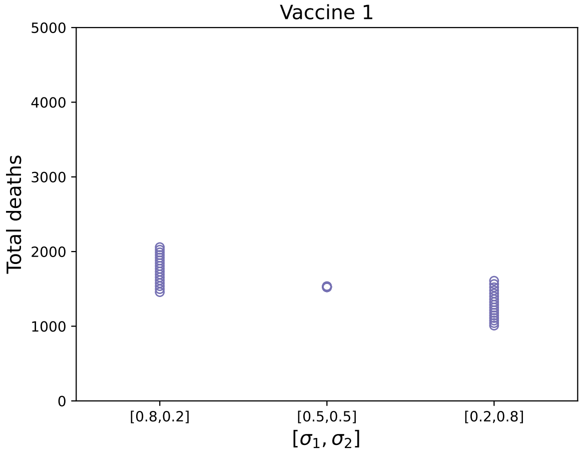


**(c)**

**Figure S10:** *Comparing total infections, symptomatic infections and deaths for Vaccine 1 as we vary* $\sigma_1$ *and* $\sigma_2$*.*

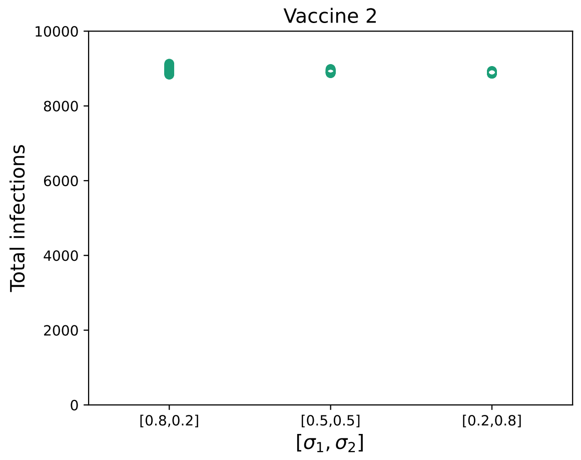


**(a)**

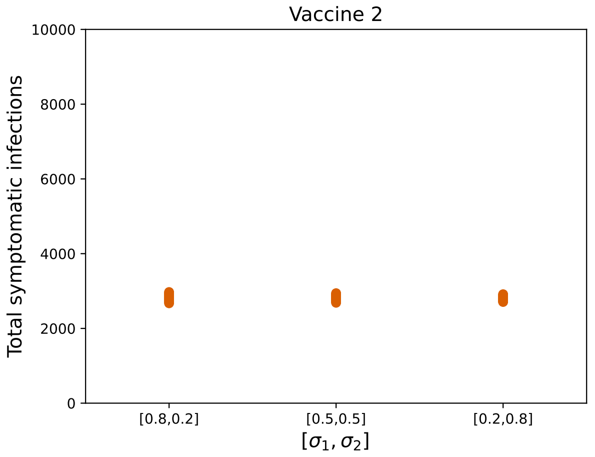


**(b)**

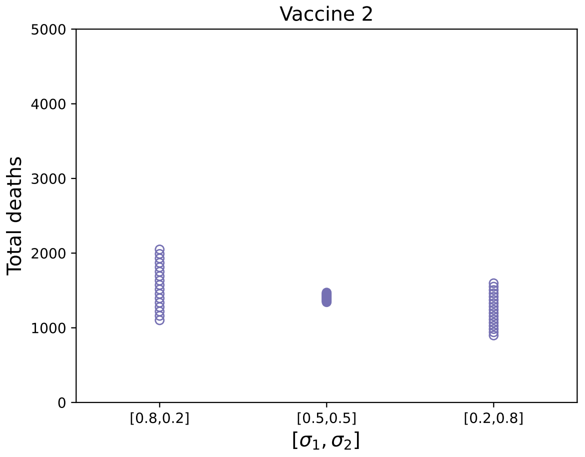


**(c)**

**Figure S11:** *Comparing total infections, symptomatic infections and deaths for Vaccine 2 as we vary* $\sigma_1$ *and* $\sigma_2$.

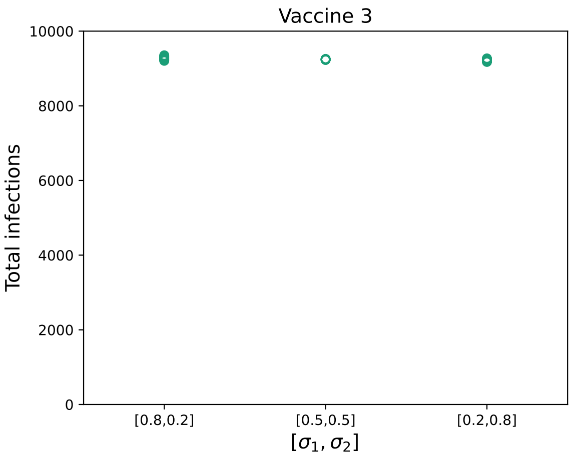


**(a)**

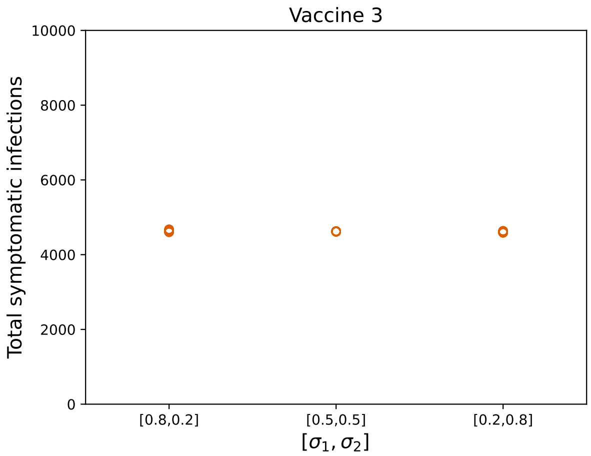


**(b)**

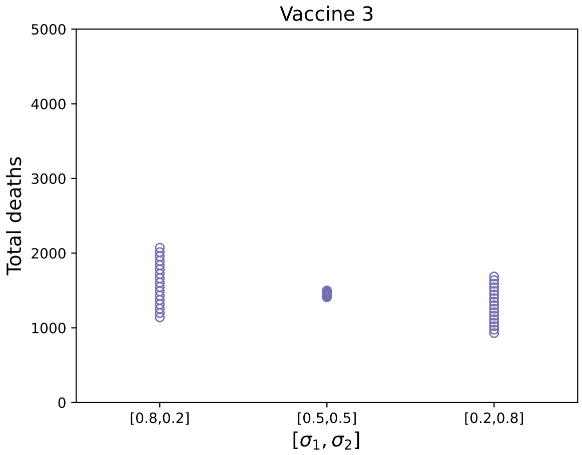


**(c)**

**Figure S12:** *Comparing total infections, symptomatic infections and deaths for Vaccine 3 as we vary* $\sigma_1$ *and* $\sigma_2$*.*

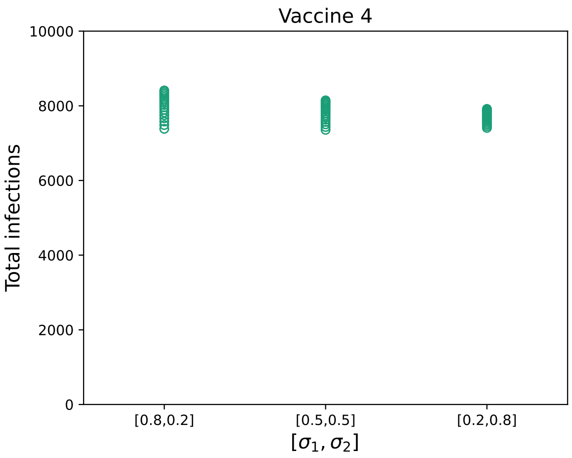


**(a)**

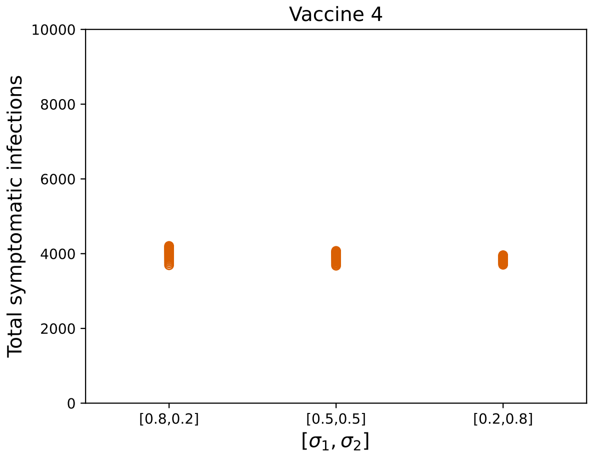


**(b)**

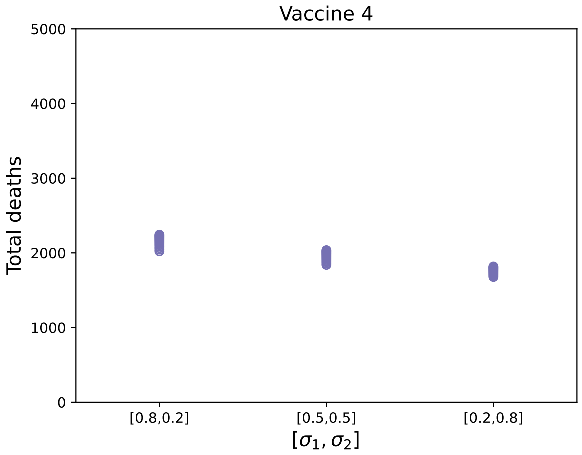


**(c)**

**Figure S13:** *Comparing total infections, symptomatic infections and deaths for Vaccine 4 as we vary $\sigma_1$ and $\sigma_2$.*

#### S2.2.4 Varying $\delta_A$

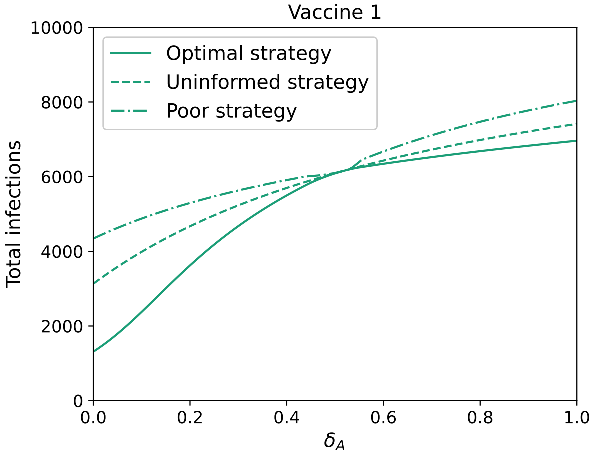


**(a)**

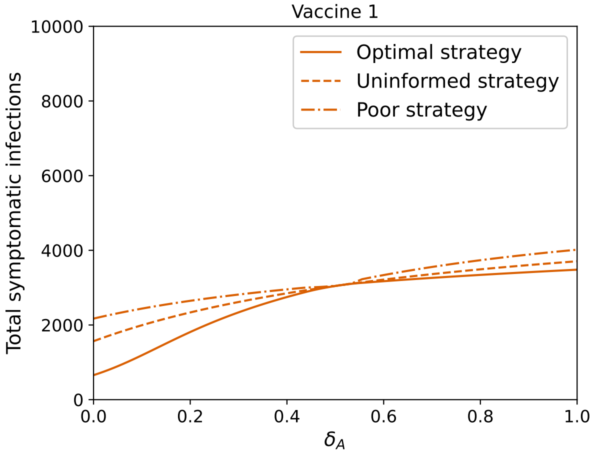


**(b)**

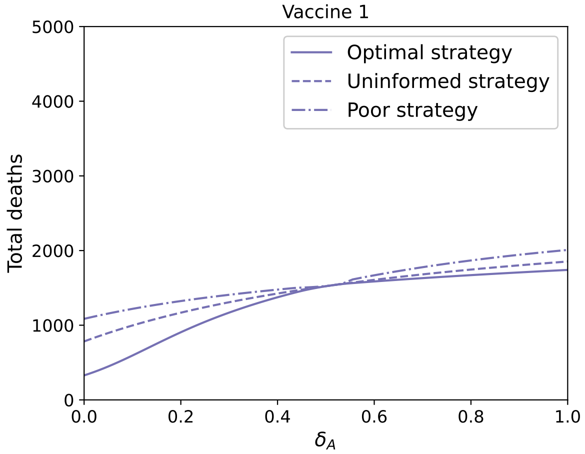


**(c)**

**Figure S14:** *Comparing total infections, symptomatic infections and deaths for Vaccine 1 as we vary* $\delta_A$.

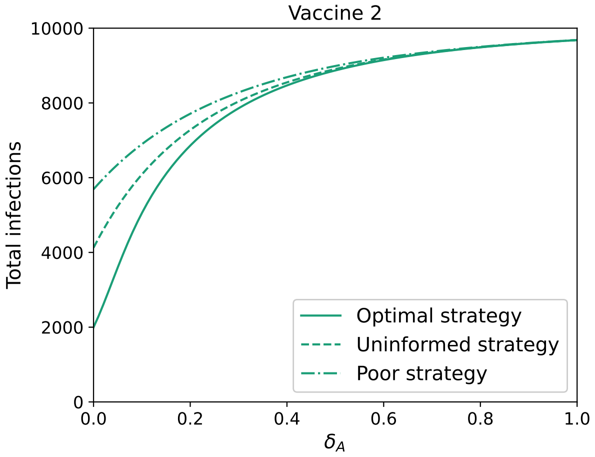


**(a)**

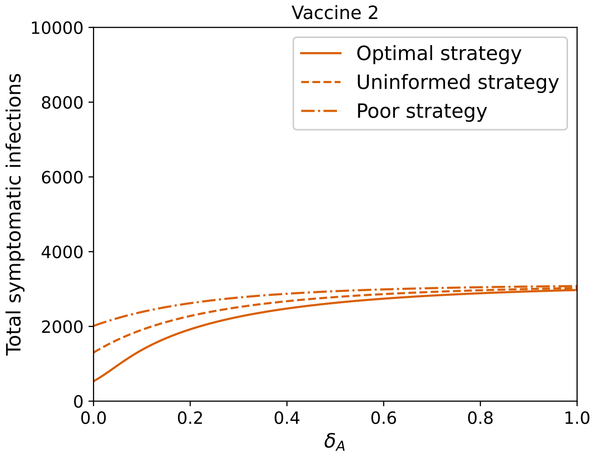


**(b)**

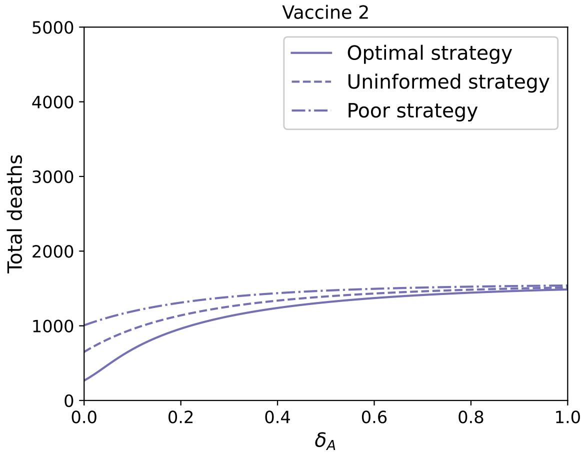


**(c)**

**Figure S15:** *Comparing total infections, symptomatic infections and deaths for Vaccine 2 as we vary* $\delta_A$.

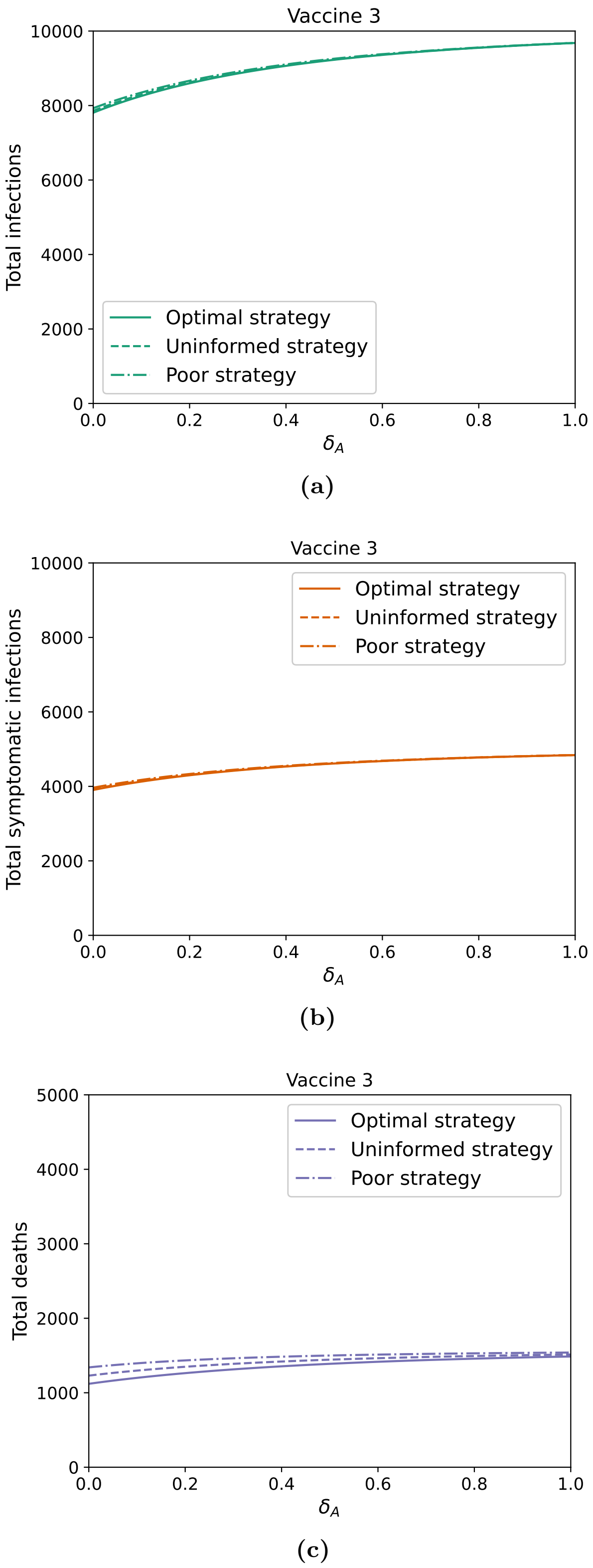


**Figure S16:** *Comparing total infections, symptomatic infections and deaths for Vaccine 3 as we vary* $\delta_A$.

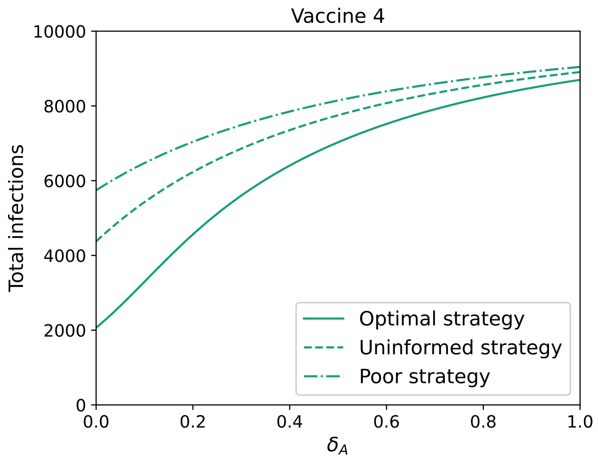


**(a)**

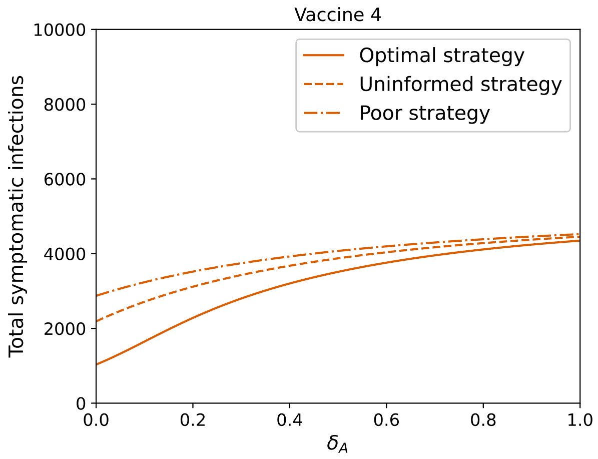


**(b)**

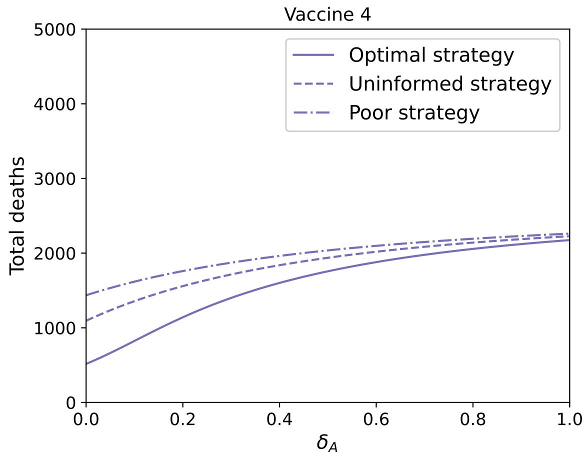


**(c)**

**Figure S17:** *Comparing total infections, symptomatic infections and deaths for Vaccine 4 as we vary* $\delta_A$.

#### S2.2.5 Varying $R_0$

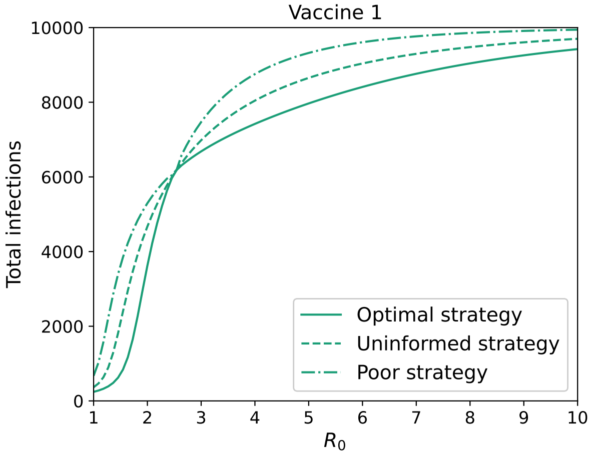


**(a)**

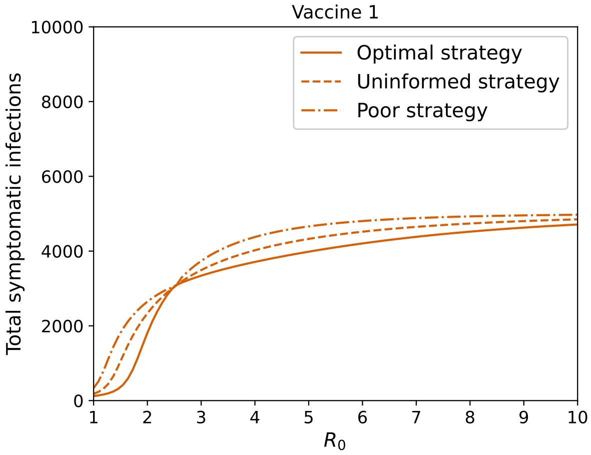


**(b)**

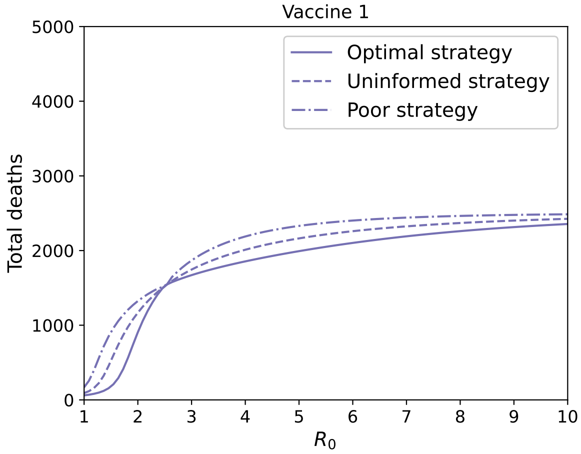


**(c)**

**Figure S18:** *Comparing total infections, symptomatic infections and deaths for Vaccine 1 as we vary* $R_0$.

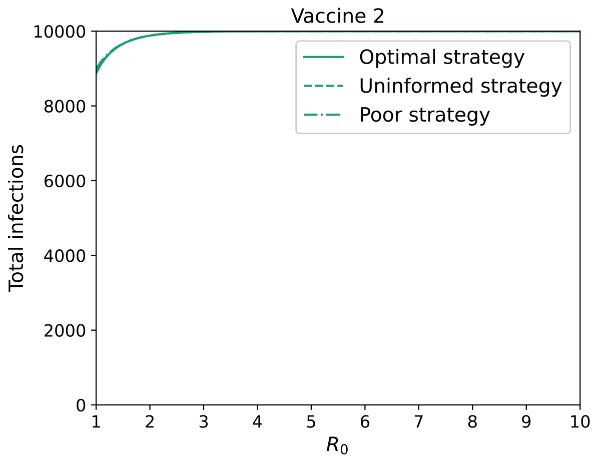


**(a)**

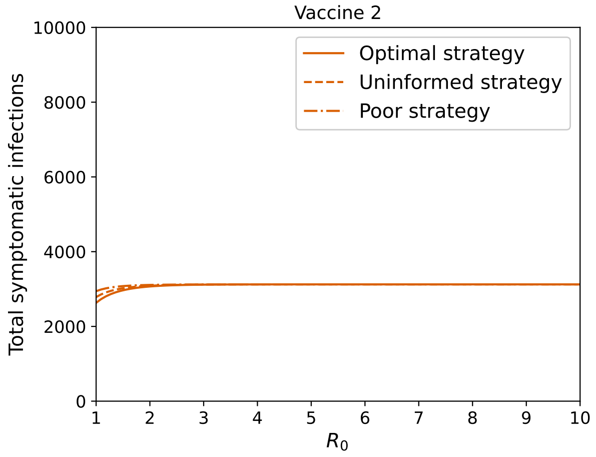


**(b)**

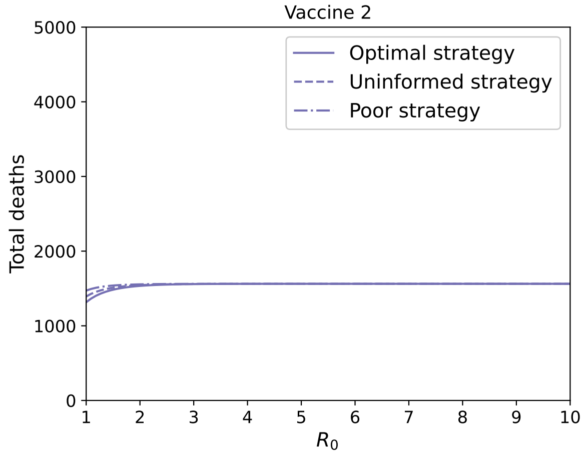


**(c)**

**Figure S19:** *Comparing total infections, symptomatic infections and deaths for Vaccine 2 as we vary* $R_0$.

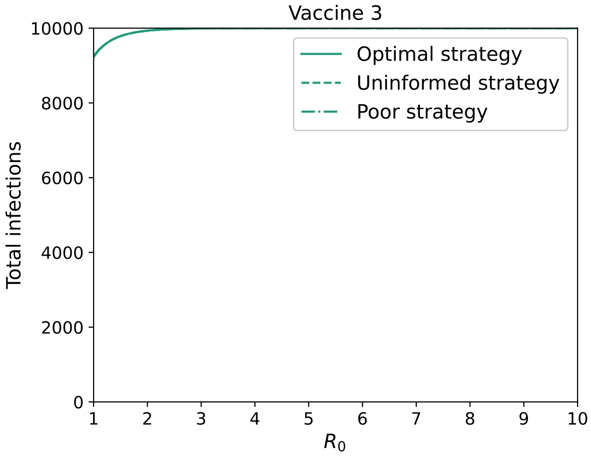


**(a)**

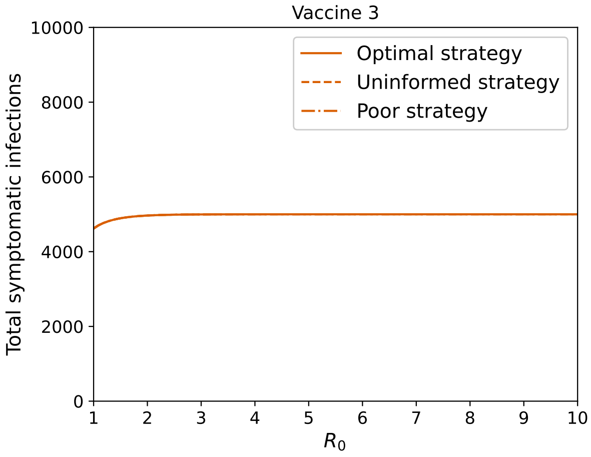


**(b)**

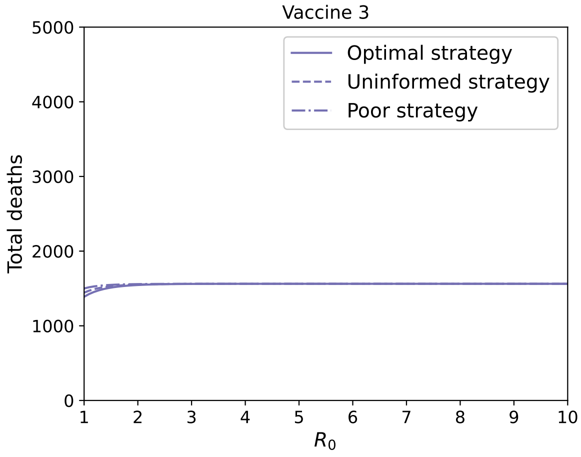


**(c)**

**Figure S20:** *Comparing total infections, symptomatic infections and deaths for Vaccine 3 as we vary $R_0$.*

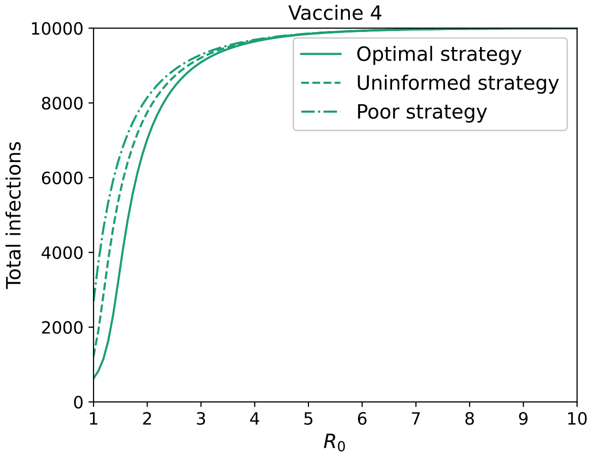


**(a)**

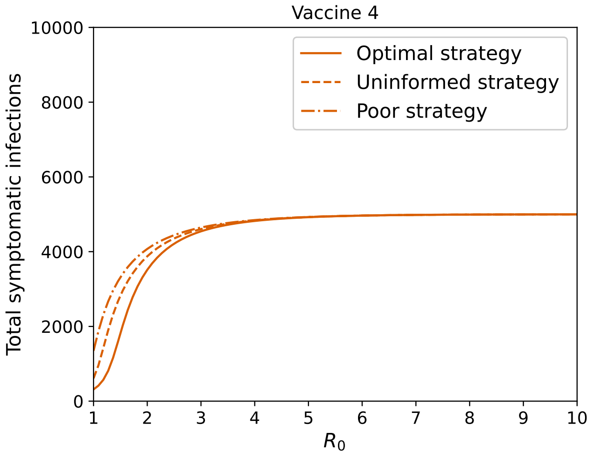


**(b)**

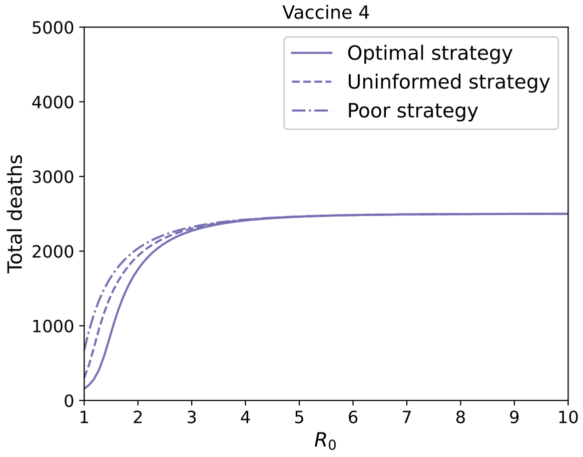


**(c)**

**Figure S21:** *Comparing total infections, symptomatic infections and deaths for Vaccine 4 as we vary* $R_0$.

#### S2.2.6 Varying vaccine coverage

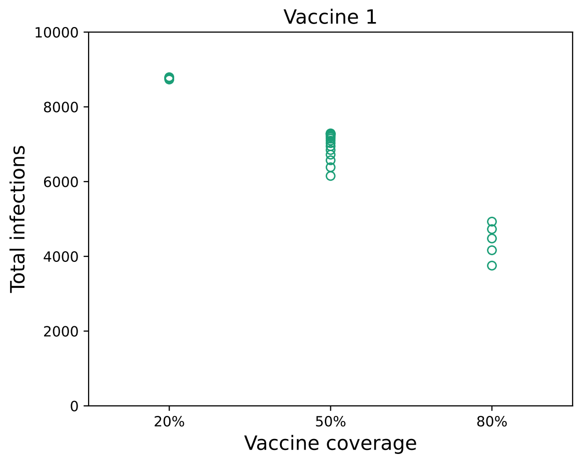


**(a)**

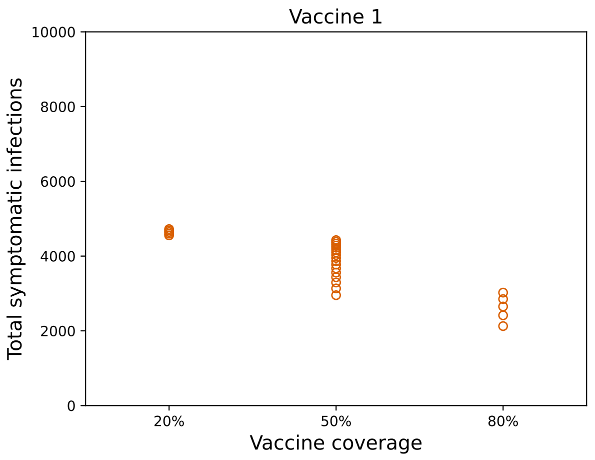


**(b)**

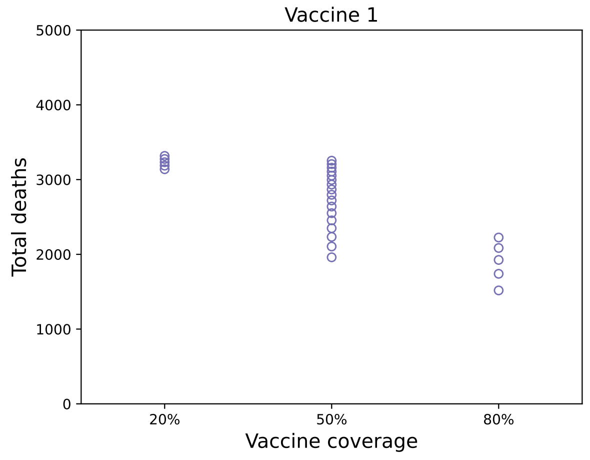


**(c)**

**Figure S22:** *Comparing total infections, symptomatic infections and deaths for Vaccine 1 as we vary vaccine coverage.*

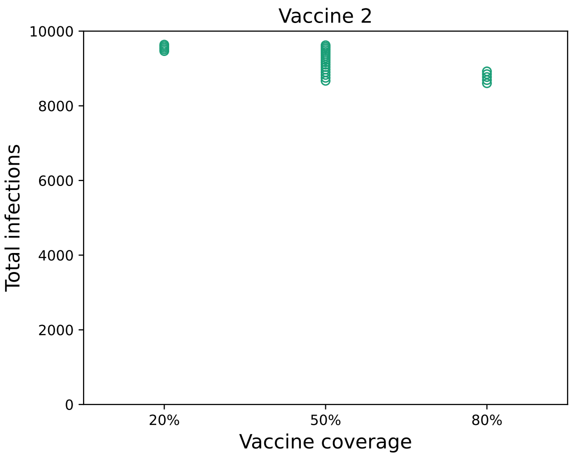


**(a)**

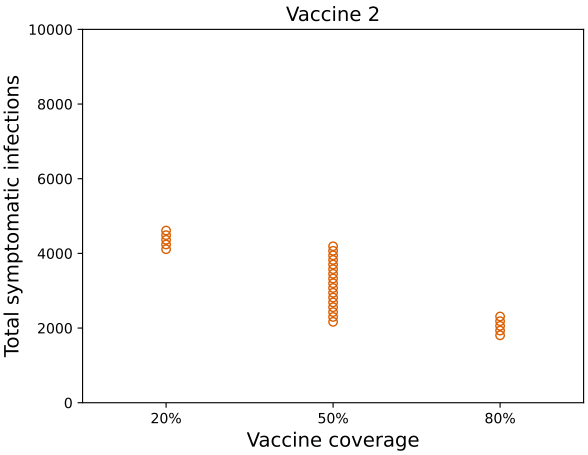


**(b)**

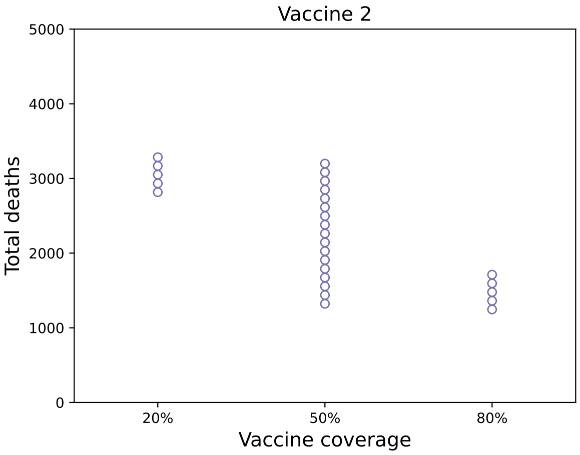


**(c)**

**Figure S23:** *Comparing total infections, symptomatic infections and deaths for Vaccine 2 as we vary vaccine coverage.*

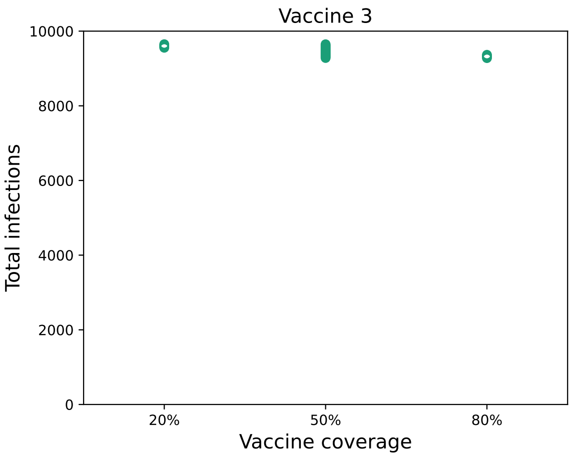


**(a)**

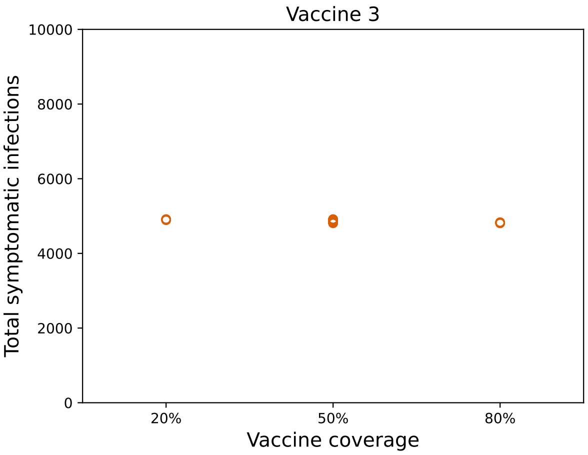


**(b)**

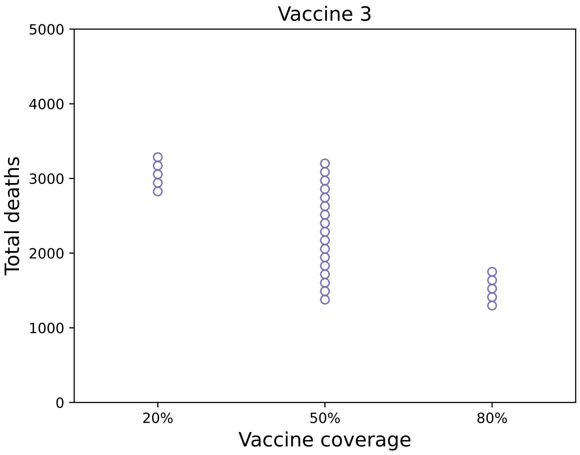


**(c)**

**Figure S24:** *Comparing total infections, symptomatic infections and deaths for Vaccine 3 as we vary vaccine coverage.*

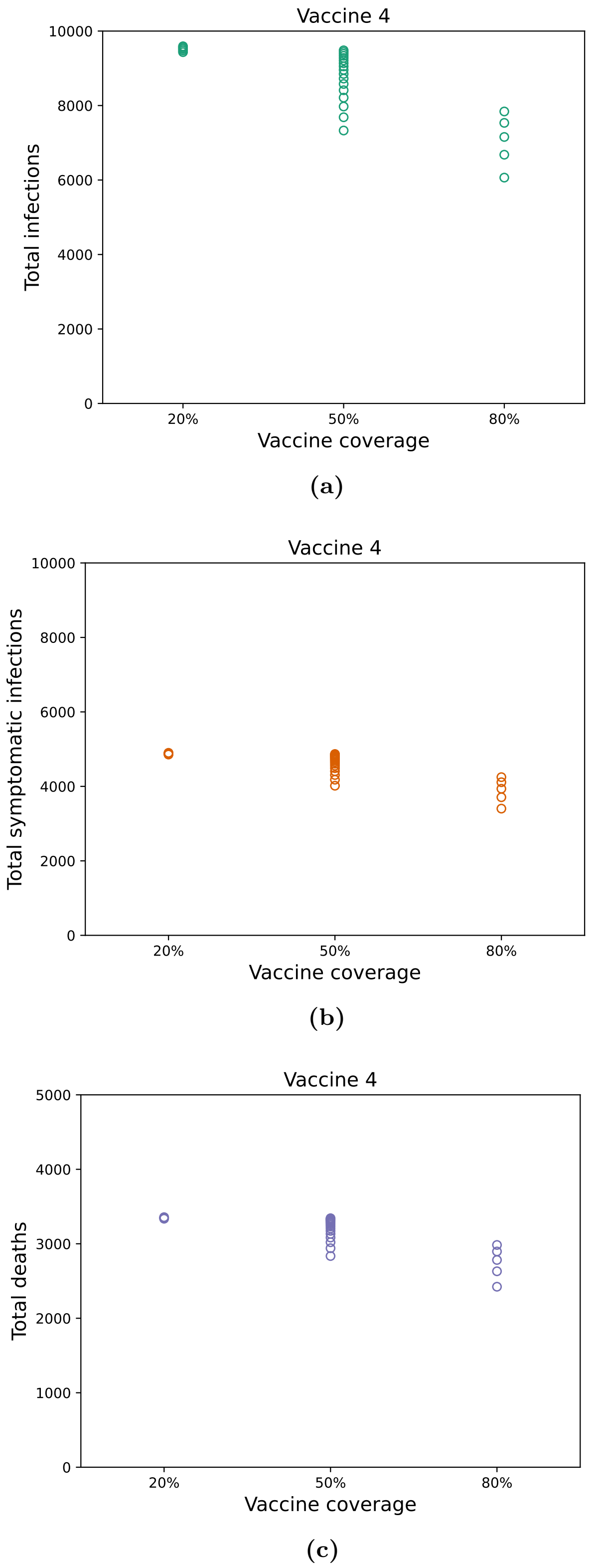


**Figure S25:** *Comparing total infections, symptomatic infections and deaths for Vaccine 4 as we vary vaccine coverage.*

#### S2.2.7 Varying vaccine effectiveness

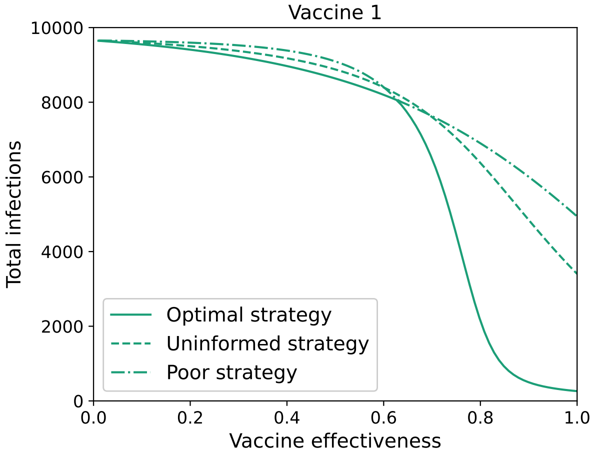


**(a)**

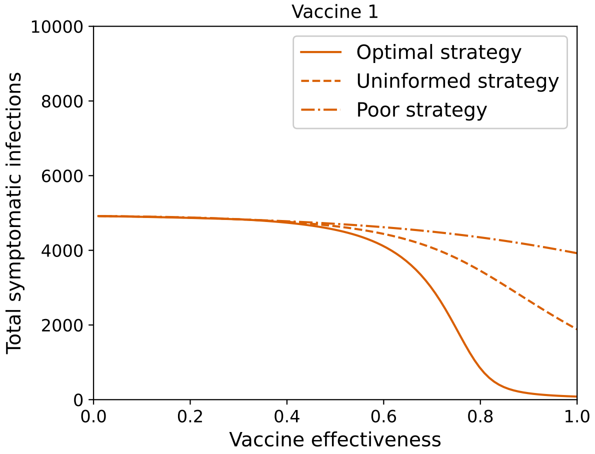


**(b)**

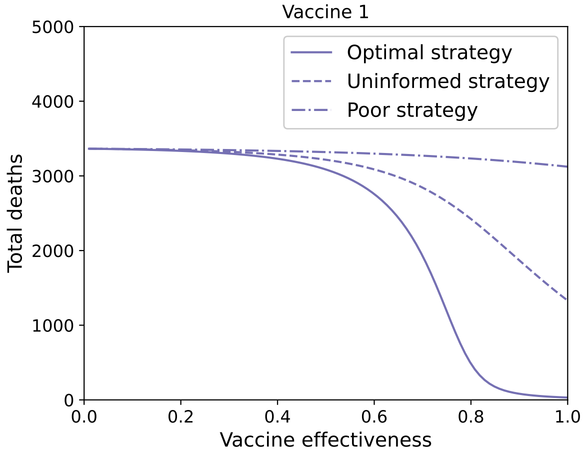


**(c)**

**Figure S26:** *Comparing total infections, symptomatic infections and deaths for Vaccine 1 as we vary vaccine effectiveness.*

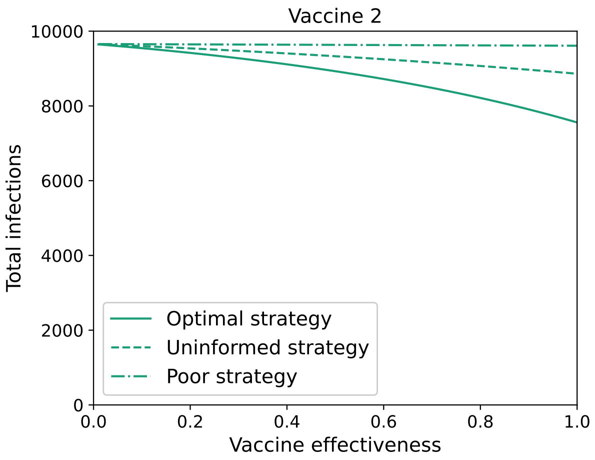


**(a)**

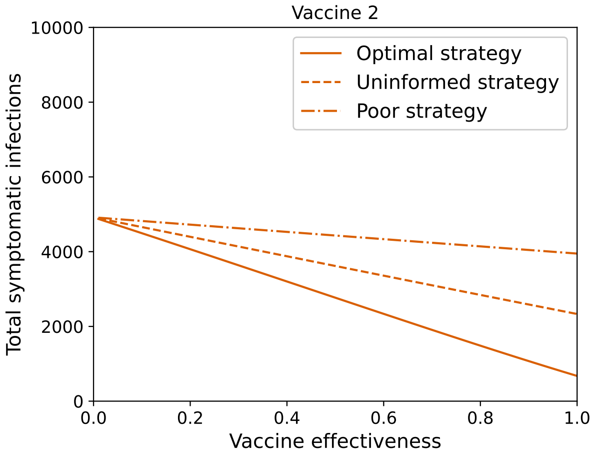


**(b)**

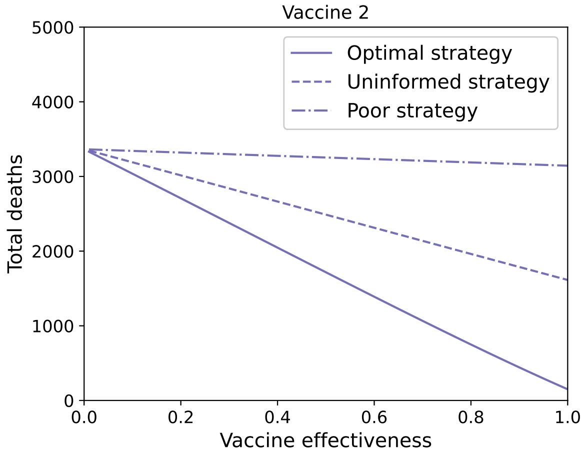


**(c)**

**Figure S27:** *Comparing total infections, symptomatic infections and deaths for Vaccine 2 as we vary vaccine effectiveness.*

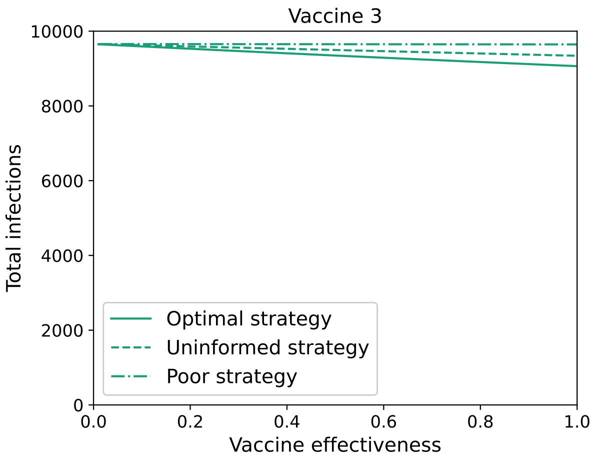


**(a)**

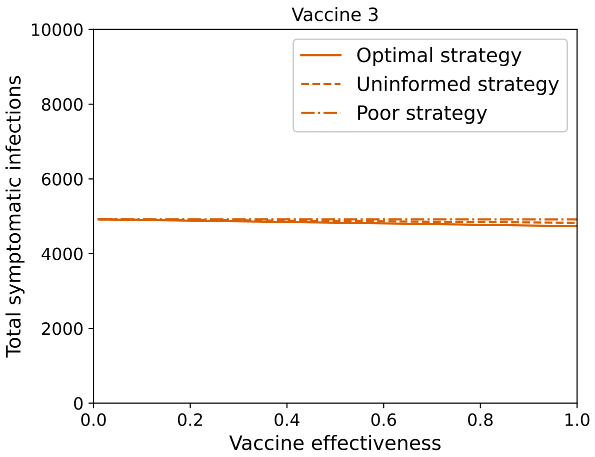


**(b)**

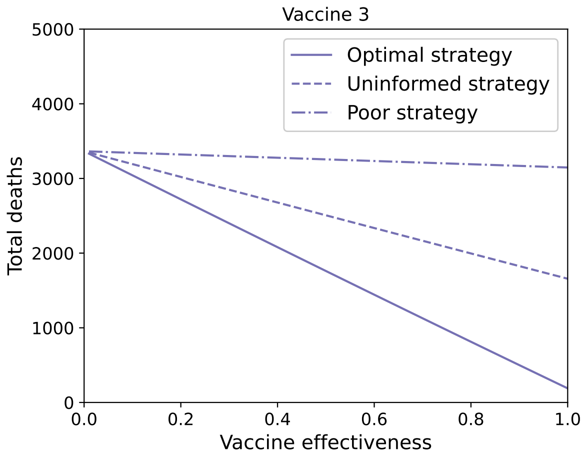


**(c)**

**Figure S28:** *Comparing total infections, symptomatic infections and deaths for Vaccine 3 as we vary vaccine effectiveness.*

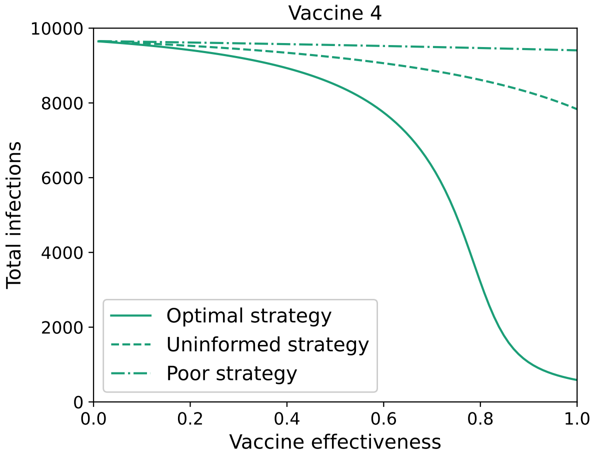


**(a)**

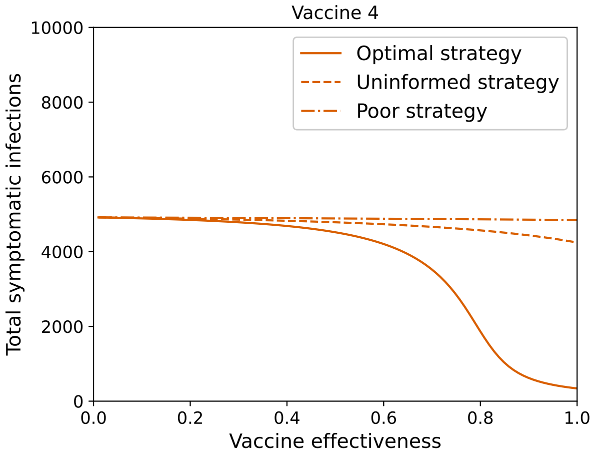


**(b)**

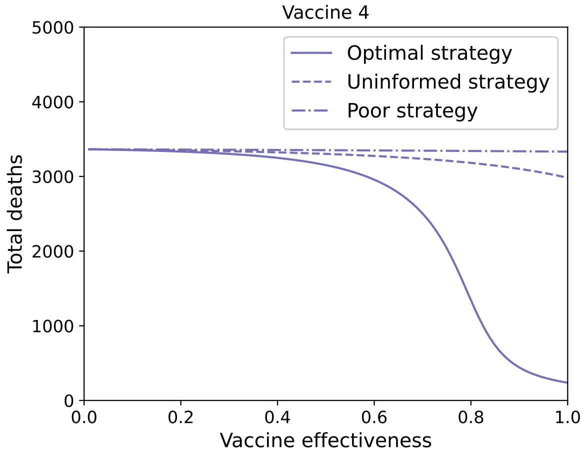


**(c)**

**Figure S29:** *Comparing total infections, symptomatic infections and deaths for Vaccine 4 as we vary vaccine effectiveness.*
